# Brain size links neurodevelopment to neurodegeneration in Parkinson’s disease

**DOI:** 10.64898/2025.12.09.25341901

**Authors:** Houman Azizi, Nooshin Abbasi, Lang Liu, Alexandre Pastor-Bernier, Christina Tremblay, Moohebat Pourmajidian, Konstantin Senkevich, Filip Morys, Andrew Vo, Roqaie Moqadam, Reza Rajimehr, Eric Yu, Peter Savadjiev, Ross D Markello, Golia Shafiei, Nina Khatibi, Neda Jahanshad, Paul M. Thompson, Jean-Baptiste Poline, Ziv Gan-Or, Bratislav Misic, Yashar Zeighami, Alain Dagher

**Affiliations:** The Neuro (Montreal Neurological Institute-Hospital), McGill University; Montreal, Quebec, H3A0G4, Canada; Douglas Mental Health University Institute, McGill University; Montreal, Quebec, H3A0G4, Canada; Department of Radiology, Brigham and Women’s Hospital, Harvard Medical School; Boston, MA, 02115, USA; Centre for Advanced Research in Sleep Medicine, Hôpital du Sacré-Cœur de Montréal; Montréal, Québec, H4J1C5, Canada; Faculty of Medicine, Department of Medicine, University of Montreal; Montreal, Quebec, H3T 1J4, Canada; School of Cognitive Sciences, Institute for Research in Fundamental Sciences (IPM); Tehran, 19395/5746, Iran; Department of Human Genetics, McGill University; Montreal, Quebec, H3A0G4, Canada; School of Computer Science, McGill University; Montreal, Quebec, H3A0G4, Canada; Department of Radiology, University of Vermont; Burlington, VT, 05405, USA; Penn Lifespan Informatics and Neuroimaging Center (PennLINC), Department of Psychiatry, Perelman School of Medicine, University of Pennsylvania; Philadelphia, PA, 19104, USA; Lifespan Brain Institute (LiBI), Children’s Hospital of Philadelphia and Penn Medicine; Philadelphia, PA, 19104, USA; Department of Biomedical Engineering, North Tehran Azad University; Tehran, 19395/1495, Iran; Imaging Genetics Center, Mark and Mary Stevens Neuroimaging and Informatics Institute, Keck School of Medicine, University of Southern California; Marina del Rey, CA, 90292, USA; Department of Neurology and Neurosurgery, McGill University; Montreal, Quebec, H3A0G4, Canada

## Abstract

Genetic studies have advanced our understanding of Parkinson’s disease (PD) pathogenesis, establishing a role for autophagy and lysosomal and mitochondrial dysfunction. However, how genetic risk translates into neuronal vulnerability remains mostly unknown. Using recent genome-wide association studies and neuroimaging data from 25,000 participants (age 44 to 85) in UK Biobank, we show that higher polygenic risk score (PRS) of PD correlates with greater cortical surface area, white matter fractional anisotropy and subcortical volumes. Mendelian randomization supports a causal relation from increased brain size to PD, and cortical regions showing the greatest polygenic expansion in surface area show the greatest atrophy in PD. Consistent with a genetically mediated increase in cortical surface area, we also find that PD polygenic risk score correlates with greater educational attainment, socioeconomic status, and other cognitive traits. Lifespan gene expression and pathway-specific analyses identify mitochondrial, autophagy, lysosomal, and neurodevelopmental pathways as separate mechanisms of genetic vulnerability. We show that a portion of PD susceptibility originates from neurodevelopmental processes that regulate neuronal proliferation. In further support of a neurodevelopmental mechanism of genetic risk, we also observed an association between PD PRS and increased cortical and subcortical size in 9-11 year olds from the ABCD cohort. Genetically-determined increases in brain size confer vulnerability to PD in later life, supporting the existence of shared neurobiological pathways between brain development and neurodegeneration.

## Introduction

Parkinson’s Disease (PD) is a progressive neurodegenerative disorder that usually presents after midlife ^1^ and is associated with a diffuse pattern of brain atrophy on magnetic resonance imaging (MRI) ^2–6^. Despite recent advances in understanding PD pathology, an unresolved question is what makes certain individuals and neural systems vulnerable to the disease. Human genetic studies have begun to address this question. Until recently, PD was thought to consist of either rare monogenic forms or sporadic cases of environmental origin, but it is now clear that genetic influences are also at play in sporadic cases. Genome-wide association studies (GWAS) have identified numerous common variants with small cumulative effects that, when aggregated into a polygenic risk score (PRS), explain up to 36% of PD heritability ^7^. Genes identified from familial cases and GWAS convergently point to specific biological pathways: protein homeostasis, alpha-synuclein misfolding and accumulation, autophagy and lysosomal dysfunction, mitochondrial impairment, and neuroinflammation ^8–10^. For example, mutations in key genes implicated in familial and sporadic PD including *SNCA* (encoding alpha-synuclein) and *LRRK2* (regulating lysosomal function and alpha-synuclein toxicity) ^11^ cause autosomal dominant PD, while *GBA1* mutations (encoding a lysosomal enzyme involved in alpha-synuclein degradation) ^9^ confer increased risk ^12,13^. However, the biological consequences of most of the loci identified by GWAS remain poorly understood.

The recent availability of large databases combining individual genetic, neuroimaging, clinical and behavioral information now make it possible to measure the effects of genetic risk in asymptomatic individuals. Leveraging GWAS, individual PRSs can be computed to identify the contribution of gene variants to brain structure and function, and to relate these to individual phenotypes of interest. Such analyses provide an opportunity to identify preclinical measures of disease vulnerability and gain a better understanding of the neurodegenerative process. Here we used data from the UK Biobank ^14^ to investigate how genetic risk for PD shapes brain anatomy and behavior. We combined genetic, neuroimaging, and behavioral data to uncover the neural and clinical correlates of the genetically determined high-risk state. We predicted that PD-PRS would be associated with evidence of subclinical neurodegeneration, such as reduced cortical and subcortical volume. Instead, we found that PD-PRS was correlated with widespread increases in cortical surface area, white matter fractional anisotropy and volumes of subcortical nuclei. These features together are thought to reflect increased neural progenitor divisions in the prenatal period, leading to increased numbers of cortical units and subcortical projections. PD-PRS was also associated with higher education and socioeconomic measures. These results suggest that genetic vulnerability to synucleinopathy may paradoxically emerge from neurodevelopmental factors that promote brain growth, possibly conferring early life advantages but increasing vulnerability to neurodegeneration in later life.

## Materials and Methods

### Participants

We used data from the UK Biobank (UKB), a prospective study of over 500,000 participants aged 37–73 at baseline (2006–2010) ^14^, under UKB approvals #35605 (PI Dagher) and #45551 (PI AC Evans). Participants provided written informed consent (http://biobank.ctsu.ox.ac.uk/crystal/field.cgi?id=200).

Participants were excluded based on: history of any neurological disorder, body mass index (BMI) > 35, relatedness closer than cousin, genetic and self-reported sex mismatch, and non-European genetic ancestry. Participants with first-degree family history of PD were also excluded (N=980) since they were used as proxy cases in PD GWAS calculation (Supplementary Table 1). Participants with brain measures beyond 3 standard deviations in each modality were further excluded. The neuroimaging dataset included up to 46,349 participants with at least 23,245 individuals (10,756 males; 12,489 females) present in each modality after exclusions and quality control. Detailed exclusion criteria and demographics are presented in Supplementary Methods and Supplementary Table 1. Open-access data from other sources used here derive from studies approved by the relevant local ethics boards.

### Brain imaging

All participants were scanned on a 3T Siemens Skyra at one of three UK sites ^15,16^. We analyzed six modalities: cortical thickness and surface area, subcortical volumes, white matter fractional anisotropy (FA) and mean diffusivity (MD), and quantitative susceptibility mapping (QSM) of 16 subcortical nuclei ^17^.

T1-weighted images were processed in-house using FreeSurfer v7.4.1 ^18,19^, with cortical parcellation based on the Desikan Killiany Tourville (DKT) atlas ^20,21^ and subcortical segmentation based on the Harvard-Oxford atlas ^22^. All outputs were visually inspected and quality controlled. Diffusion MRI (dMRI) images were processed using TractoFlow v2.2.1 ^23^ to generate FA and MD maps. Subject-specific diffusion brain maps and the white matter anatomical tract atlases were registered to the ICBM 152 nonlinear 2009c atlas ^24^, and diffusion measures were averaged across 73 anatomical tracts derived from the O’Donnell Research Group (ORG) atlas ^25^. QSM data were obtained from UKB for subcortical nuclei from the Harvard-Oxford subcortical atlas, as well as the substantia nigra (16 nuclei) ^17^. Acquisition parameters, preprocessing pipelines, and quality control are described in Supplementary Methods. The diffusion pipeline is available at https://github.com/HoumanAzizi/UKB_DTI_Pipeline.

### Pathway gene selection

To assess pathway-specific genetic risk of PD, we curated three gene sets for key cellular processes implicated in PD pathogenesis: mitochondrial function (1,997 genes), lysosomal function (501 genes), and autophagy (428 genes), derived from MitoCarta3.0 ^26,27^, Gene Ontology ^28,29^, and the Human Autophagy Database (collectively termed Mito-ALP pathways; Supplementary Methods). Single-nucleotide polymorphisms (SNPs) that did not fall within these genes were labelled non-Mito-ALP SNPs (see Supplementary Table 2 for N_SNP_ per pathway).

### PD-PRS calculation

Polygenic risk scores for PD (PD-PRS) were calculated using the PRS-CS method ^30^ and PLINK 2.0 ^31,32^ with the “auto” global shrinkage parameter. Posterior SNP weights were inferred using the latest PD GWAS summary statistics ^33^ as the discovery dataset and the 1000 Genomes European sample as the LD reference panel ^34^. Pathway-specific PD-PRS were calculated using the same approach restricted to SNPs within genes defined in the previous section. Given that pathway-specific PD-PRSs with fewer SNPs may have reduced statistical power, we constructed an additional PD-PRS using the top 687,574 non-Mito-ALP SNPs (similar N SNPs to the combined Mito-ALP PD-PRS) to evaluate whether pathway size alone could account for potential associations. As a sensitivity analysis, polygenic risk scores were also computed and analyses were repeated using the PRSice-2 method ^35^. Details of genetic quality control and PD-PRS calculation method for each dataset are further described in Supplementary Methods.

### Linear regression models

We performed linear regression to localize the spatial relationship between PD-PRS and the following brain measures: cortical thickness and surface area, subcortical volume, subcortical QSM, and white matter FA and MD. All models included the following covariates: age, age squared, sex, sex:age interactions, number of days since start of image acquisition, number of days squared, average head motion during functional MRI, table position (z-axis) inside the scanner, and scan site. Genotyping batch, and the top 15 genetic principal components (PCs) were further accounted for in analyses including genetic data ^36,37^. Total intracranial volume (TIV) is inherently correlated with our measures of interest such as surface area and subcortical volume, and was therefore not included as a covariate to avoid removing neurobiologically meaningful variance from the models. P-values were corrected for multiple comparisons across brain regions or tracts using the false discovery rate (FDR) approach with an FDR-corrected p-value of 0.05 as significance threshold ^38,39^. To examine the sex-specific effects of PD-PRS, we further repeated similar analyses in males and females separately and assessed spatial similarity of PD-PRS cortical effect maps in males and females using spin-test ^40,41^.

To quantify the unique contribution of PD-PRS in each model, we computed partial R²_PD-PRS_ representing the proportion of residual variance explained by PD-PRS after accounting for all covariates, using the following formula: (R²_complete_model_ - R²_reduced_model_) / (1 - R²_reduced_model_), where the reduced model contained only the covariates.

Additionally, to replicate the neuroanatomical associations of PD-PRS in an adolescent developmental cohort, we repeated the regression analyses in children and adolescents from the Adolescent Brain Cognitive Development (ABCD) cohort ^42–44^ across four longitudinal time points (ages 10 to 16 years old). See Supplementary Methods for ABCD sample characteristics and details.

### Spatial correlation analyses

We compared the spatial patterns of PD-PRS cortical associations against PD-related cortical atrophy and surface area maps from the ENIGMA-PD consortium, which compared 3,096 PD patients across Hoehn and Yahr stages to healthy controls while accounting for age and sex ^45^. Statistical significance was assessed using spin-test spatial null models to account for spatial autocorrelation among brain regions ^40,41^ (see Supplementary Methods for details).

### PD-PRS and behavioral characteristics

We tested the association of PD-PRS with behavioral phenotypes relevant to PD in healthy UKB participants. Behavioral phenotypes included cognitive measures (education level, fluid intelligence), socioeconomic indicators (household income, multiple deprivation index), lifestyle factors (coffee intake, sleep duration, BMI, smoking, alcohol usage, addiction), and potential prodromal PD features (hand grip strength, constipation, orthostatic hypotension, urinary incontinence, apathy and depressed mood). See Supplementary Methods for full trait definitions. All behavioral variables were z-score normalized. Analyses were performed using linear regressions (FDR-corrected) while controlling for age, age squared, sex:age interaction, study site, genotyping batch and the top 15 population genetic PCs ^36^. Sample sizes varied between 29,890 and 111,328 participants (Supplementary Table 3).

### Mendelian randomization

Two-sample Mendelian randomization (MR) was used to test whether whole-brain and region-wise neuroanatomical measures have a causal effect on PD diagnosis. Exposures were GWAS summary statistics for whole-brain total cortical surface area and average thickness ^46^, total subcortical grey matter volume ^47^, and average white matter FA ^48^, as well as region-wise cortical surface area, cortical thickness, subcortical volume, subcortical QSM, and white matter FA ^17,47^. The latest PD GWAS summary statistics were used as the outcome ^33^. Analyses were performed using the TwoSampleMR package in R ^49,50^ using inverse-variance weighted (IVW), MR Egger, and weighted median methods ^51–54^ while controlling for heterogeneity and horizontal pleiotropy ^55,56^. Potential reverse MR and the power achieved to detect an effect size equivalent to the odds ratio of 1.2 of PD diagnosis with 95% confidence were further calculated (see Supplementary Methods for MR parameters) ^57^.

### Gene expression analysis

PD risk genes were defined as those prioritized by Yu et al. ^58^ based on the Nalls et al. 2019 PD GWAS ^7^ with probability score over 0.5, as well as novel genes identified by the latest PD GWAS ^33^. PD genes were partitioned into four pathways: lysosomal (n = 17), mitochondrial (n = 29), autophagy (n = 7), and non-Mito-ALP PD risk genes (n = 214; Supplementary Table 4). The developmental expression trajectories of each PD risk gene set from pre-natal stages to adulthood was then assessed using the BrainSpan RNA-seq dataset ^59^. The average expression trajectory across all genes in the BrainSpan dataset was also calculated as a reference. Calculation and normalization steps of the BrainSpan data are detailed in Supplementary Methods.

### Multi-marker Analysis of GenoMic Annotation (*MAGMA*)

Gene-level analysis was performed using MAGMA v1.08 through the FUMA platform v1.8.0 ^60,61^, separately for Mito-ALP and non-Mito-ALP SNP subsets of the latest PD GWAS ^33^. Gene-set enrichment was subsequently assessed in over 15,000 gene sets from the Molecular Signatures Database (MSigDB v2023.1.Hs) ^62^ using MAGMA’s competitive gene-set analysis. Multiple comparisons were corrected using the Bonferroni method (see Supplementary Methods for detailed parameters).

### Linkage disequilibrium score regression (LDSC) analysis

Genetic correlations between PD (full and pathway-specific GWAS) ^33^ and cortical thickness, cortical surface area, and subcortical volumes ^63,64^ were calculated using LDSC ^65^ to determine whether shared genetic influences contributed to both PD and brain structure. FDR correction was applied across brain regions.

## Results

### Sample characteristics

To identify the effects of PD genetic risk on the brain, we studied MRI, genetic, and behavioral data of 46,349 healthy participants aged 44 to 85 years old (mean 65.1; before exclusions) from the UKB ^14,15^. After exclusions and quality control (see Methods), our final neuroimaging dataset included up to 46,349 healthy UKB participants aged 44 to 85 years (mean 65.1), with at least 23,245 individuals (10,756 males; 12,489 females) present in each imaging modality. Sample sizes for each analysis are summarized in Supplementary Table 5. Characteristics for the ABCD dataset are described below.

### Genetic risk for PD differentially impacts cortical surface expansion and thickness

We fitted general linear regression models between PD-PRS and neuroanatomical measures while adjusting for potential covariates and correcting for multiple comparisons using the FDR approach (see Methods). PD-PRS showed a significant positive association with average surface area (Figure 1a; p_FDR_<0.001; sample size N=25,256) and with surface area in all individual cortical regions of the DKT atlas (Figure 1b; Supplementary Table 6). The effect was similar across hemispheres (r=0.86, p < 0.001) and PD-PRS explained 0.74% of average surface area variance after adjusting for covariates (partial r^2^_PD-PRS_=0.0074; Supplementary Table 7). Conversely, PD-PRS was negatively associated with global mean cortical thickness (Figure 1a; p_FDR_<0.01; N=25,237; partial r^2^_PD-PRS_=0.0004). Regionwise analysis revealed significant negative associations between PD-PRS and cortical thickness in a smaller number of regions primarily spanning the prefrontal, temporal, and limbic cortices, including bilateral orbitofrontal, rostral middle frontal, anterior cingulate, inferior temporal, fusiform, and inferior parietal areas (Figure 1c; Supplementary Table 6).

**Figure 1.**
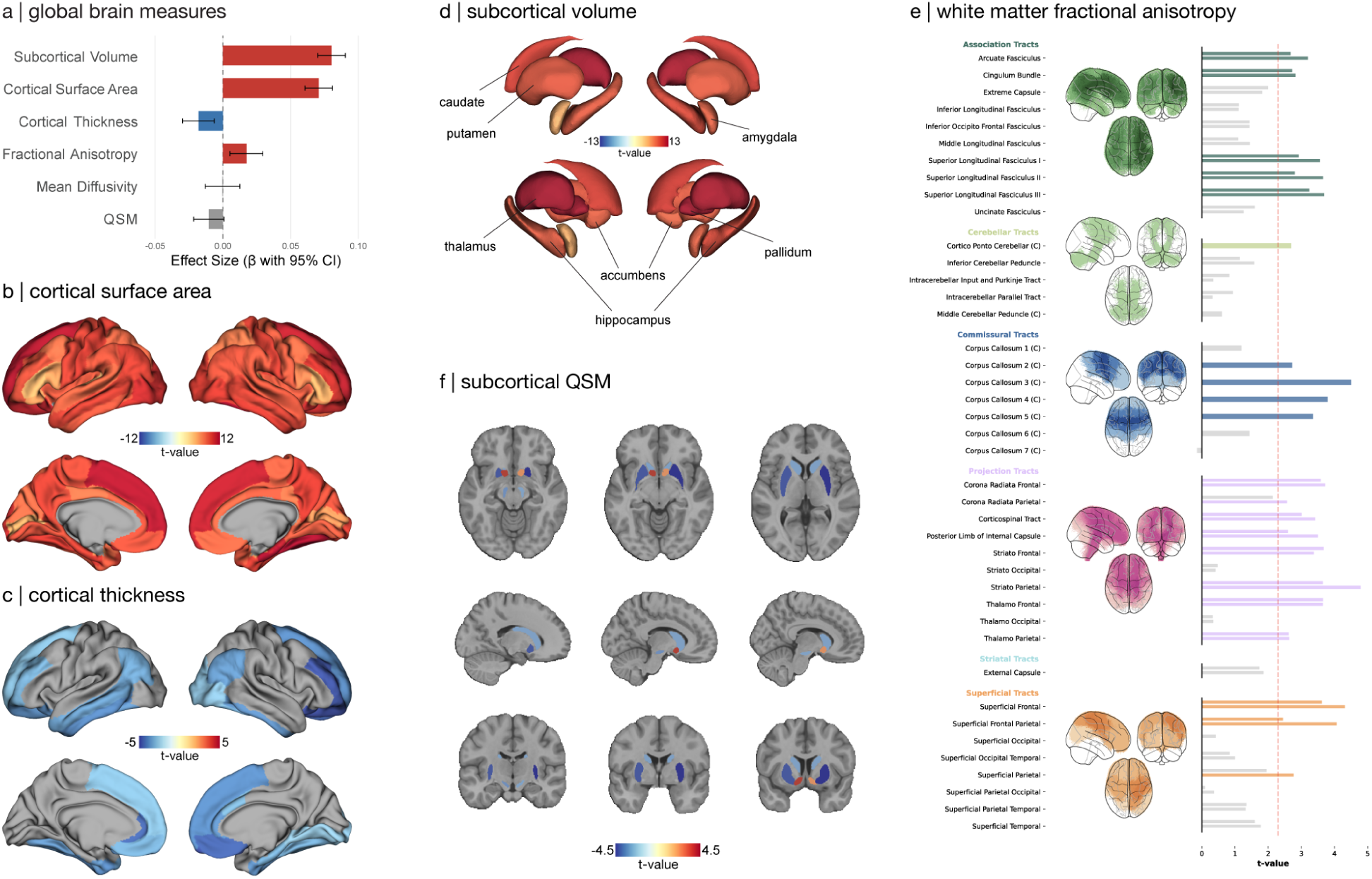
Association between PD-PRS and morphometrical brain measures. **(A)** Effect size and 95% confidence intervals of the effect of PD-PRS on global morphometrical brain measures. **(B)** T-statistics maps showing the effect of PD-PRS on cortical surface area. **(C)** T-statistics maps showing the effect of PD-PRS on cortical thickness. **(D)** T-statistics maps showing the effect of PD-PRS on subcortical volume. **(E)** Glass brain and bar plots showing t-statistics for the effect of PD-PRS on white matter tract FA. Each glass brain shows projection of significant tracts in its corresponding tract category. Bar plots are shown as a single bar for cross-hemispheric tracts and as two bars for bilateral tracts (left hemisphere: upper bar; right hemisphere: lower bar). The red dashed line shows the FDR significance threshold. **(F)** T-statistics maps showing the effect of PD-PRS on subcortical QSM. In all plots, only regions with FDR-corrected p-value < 0.05 are colored. PD-PRS: Parkinson’s disease polygenic risk score.

### Genetic risk for PD is linked to larger subcortical volumes

PD-PRS was positively associated with total subcortical volume (Figure 1a) and with the volume of all 14 individual subcortical structures, including bilateral thalamus, caudate, putamen, pallidum, hippocampus, accumbens, and amygdala (Figure 1d, Supplementary Table 6, all p_FDR_<0.001; N=25,255; partial r^2^_PD-PRS_=0.0092).

### Genetic risk for PD is associated with widespread increase in white matter fractional anisotropy

PD-PRS was positively associated with global average FA (Figure 1a; p_FDR_<0.01; N=21,815; partial r^2^_PD-PRS_=0.0003) and with FA across 35 of 73 tracts of the ORG atlas (all p_FDR_<0.05; N=23,245). These associations were widespread and included most projection and commissural tracts, several association and superficial tracts, and the cortico-ponto-cerebellar tract (Figure 1e; Supplementary Table 6). Conversely, PD-PRS did not show any significant associations with MD (Supplementary Table 6; N>22,198).

In summary, PD-PRS was associated with widespread increases in cortical surface area, white matter FA, and subcortical volume. This global increase in brain size may reflect increased proliferation of neural progenitor cells during fetal brain development, leading to a greater number of neuronal columns and axonal projections ^63,66^. We further tested this hypothesis as described below.

### Genetic risk for PD is associated with reduced magnetic susceptibility in subcortical nuclei

QSM of the substantia nigra (SN) has been proposed as a PD biomarker. It correlates with PD severity and duration and is thought to represent iron accumulation and SN neurodegeneration ^67,68^. We therefore tested whether PD-PRS was associated with subclinical or prodromal PD by examining QSM measures across 16 subcortical regions. We observed a widespread negative association between PD-PRS and QSM in bilateral substantia nigra, putamen, and caudate (p_FDR_<0.05; N=30,225; partial r^2^_PD-PRS_=0.0001; Figure 1f; Supplementary Table 6). These results suggest that the participants with high PD-PRS in this sample do not have subclinical basal ganglia degeneration, supporting that the PRS-neuroimaging associations reported here represent a vulnerability endophenotype and not early neurodegeneration.

### Comparison with Parkinson’s disease atrophy pattern

We next asked whether the morphometric effects of genetic risk for PD resembled patterns of brain atrophy in people diagnosed with PD by comparing our PD-PRS effect maps to those of the ENIGMA-PD consortium ^45^. The PD-PRS effect on surface area (PD-PRS ∼ SA) was significantly negatively correlated with ENIGMA-PD cortical thickness maps across the first three Hoehn and Yahr (HY) disease stages and the group average (r<-0.31, p_spin_<0.05), and showed a similar significant negative relationship with the ENIGMA-PD surface area map at the third disease stage (Supplementary Figure 1a; r=-0.47, p_spin_=0.003). Conversely, the PD-PRS effect on cortical thickness (PD-PRS ∼ CT) was not significantly associated with the ENIGMA-PD maps (Supplementary Table 8; Supplementary Figure 1b). These results suggest that areas with the greatest cortical surface area expansion due to genetic risk of PD are also the most vulnerable to cortical thinning in PD.

### Testing causal association between neuroanatomical measures and PD

We investigated the nature and direction of the previously found associations between brain measures and PD using two-sample Mendelian randomization (MR). The single-nucleotide polymorphisms (SNPs) used as instrumental variables (16 to 31 per analysis; mean=23) had sufficient strength (F-statistics > 36) and power (100%) to detect an effect size equivalent to an odds ratio of 1.2 for PD diagnosis (Supplementary Table 9).

We found a potentially causal positive association between total cortical surface area and PD risk. This association remained consistent after removing pleiotropic outlier SNPs (rs56323304 and rs73802707), one of which is located within the MAPT gene (Figure 2a; IVW p<0.001, weighted median p<0.001, weighted mode p<0.01, simple mode p<0.05). Similarly, we observed a positive association between total subcortical grey matter volume and PD risk (Figure 2b; Supplementary Table 9; IVW p<0.001, weighted median p<0.01). MR-Egger intercept tests showed no evidence of directional pleiotropy in any of the analyses. However, significant heterogeneity was apparent in the instrumental SNPs of most analyses (Supplementary Table 9; IVW Cochran’s Q p<0.05). Reverse MR analyses showed no evidence of a causal effect of PD on brain structure (Supplementary Table 9). These findings indicate that genetic predisposition toward larger cortical surface area and subcortical volume is significantly linked to higher risk for PD development.

**Figure 2.**
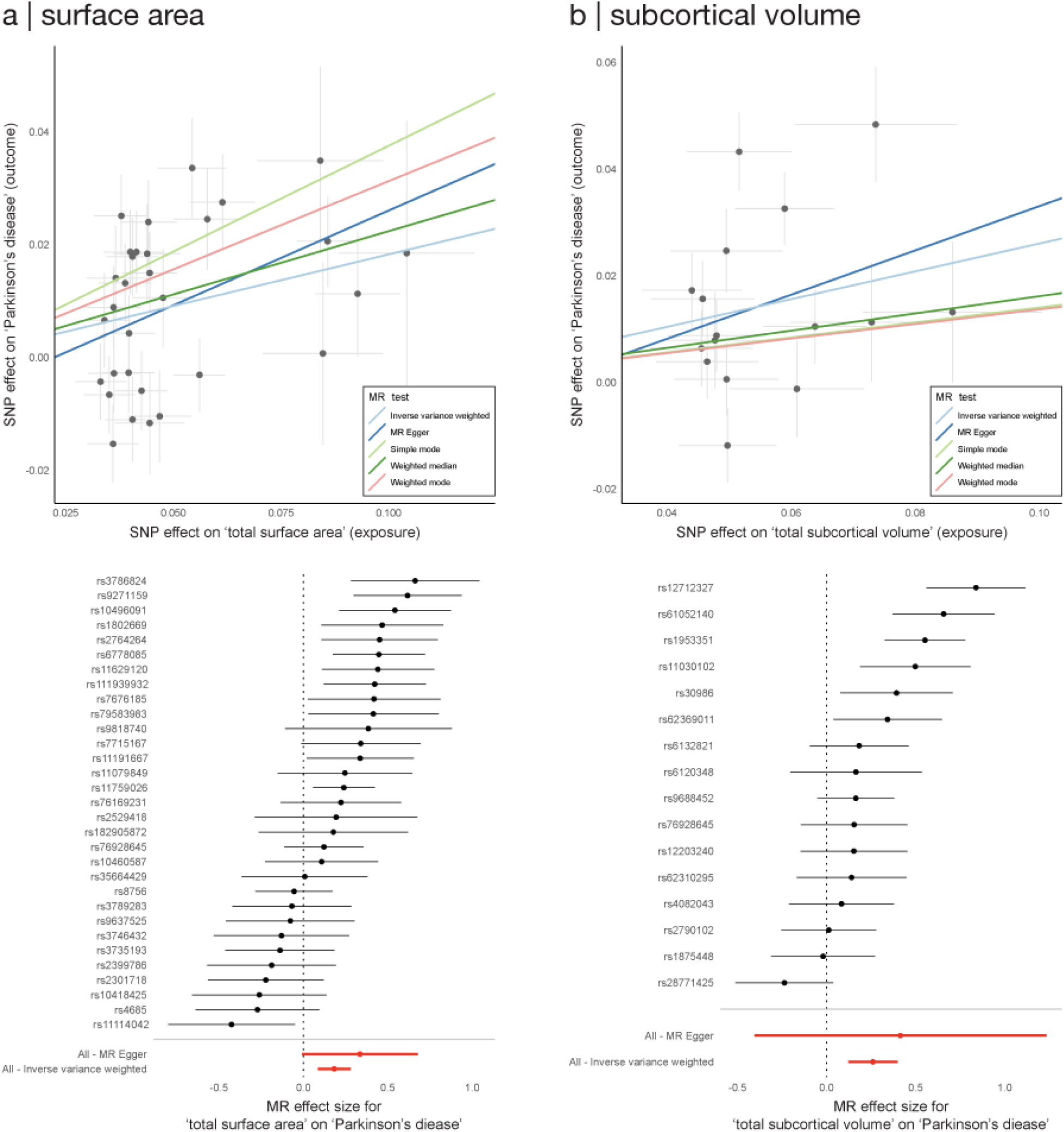
Mendelian randomization analysis of the potentially causal effects of total cortical surface area and subcortical volume on Parkinson’s disease. Scatter plots (top) and forest plots (bottom) showing Mendelian randomization results for the causal effect of **(A)** total cortical surface area and **(B)** total subcortical volume on Parkinson’s disease. Scatter plots display the effect size and 95% confidence interval of each SNP on the exposure (x-axis) and outcome (y-axis), with lines representing regression analysis using different MR methods. Forest plots show odds ratios and 95% confidence intervals for individual SNPs as well as overall estimates for MR Egger and IVW methods. MR: Mendelian randomization; IVW: inverse variance weighted.

### Behavioral correlates of genetic risk for PD

Given that genetic risk of PD was associated with brain size, we tested the relationship between PD-PRS and pertinent behavioral measures in a larger UKB sample (Figure 3a; Supplementary Table 3). PD-PRS was negatively associated with smoking, BMI, multiple deprivation index, as well as apathy and depressed mood in the past 2 weeks (all p_FDR_<0.05). Additionally, PD-PRS was positively associated with educational attainment, fluid intelligence, household income, and sleep duration (all p_FDR_<0.05). PD-PRS was associated with stronger hand grip (β=0.007; p_FDR_<0.001), further supporting the notion that the older adults with high PRS in this sample do not have subclinical PD. Coffee consumption, alcohol usage, addiction score, and clinical measures (constipation, orthostatic hypotension, and urinary incontinence) did not show significant associations (β≤0.006). Similar directions of associations were observed in the subset with brain imaging data, although only education level and household income reached statistical significance (Supplementary Figure 2; Supplementary Table 3; p_FDR_<0.05). In summary, higher PD-PRS is associated with numerous behavioral and demographic markers of cognition and socioeconomic status.

**Figure 3.**
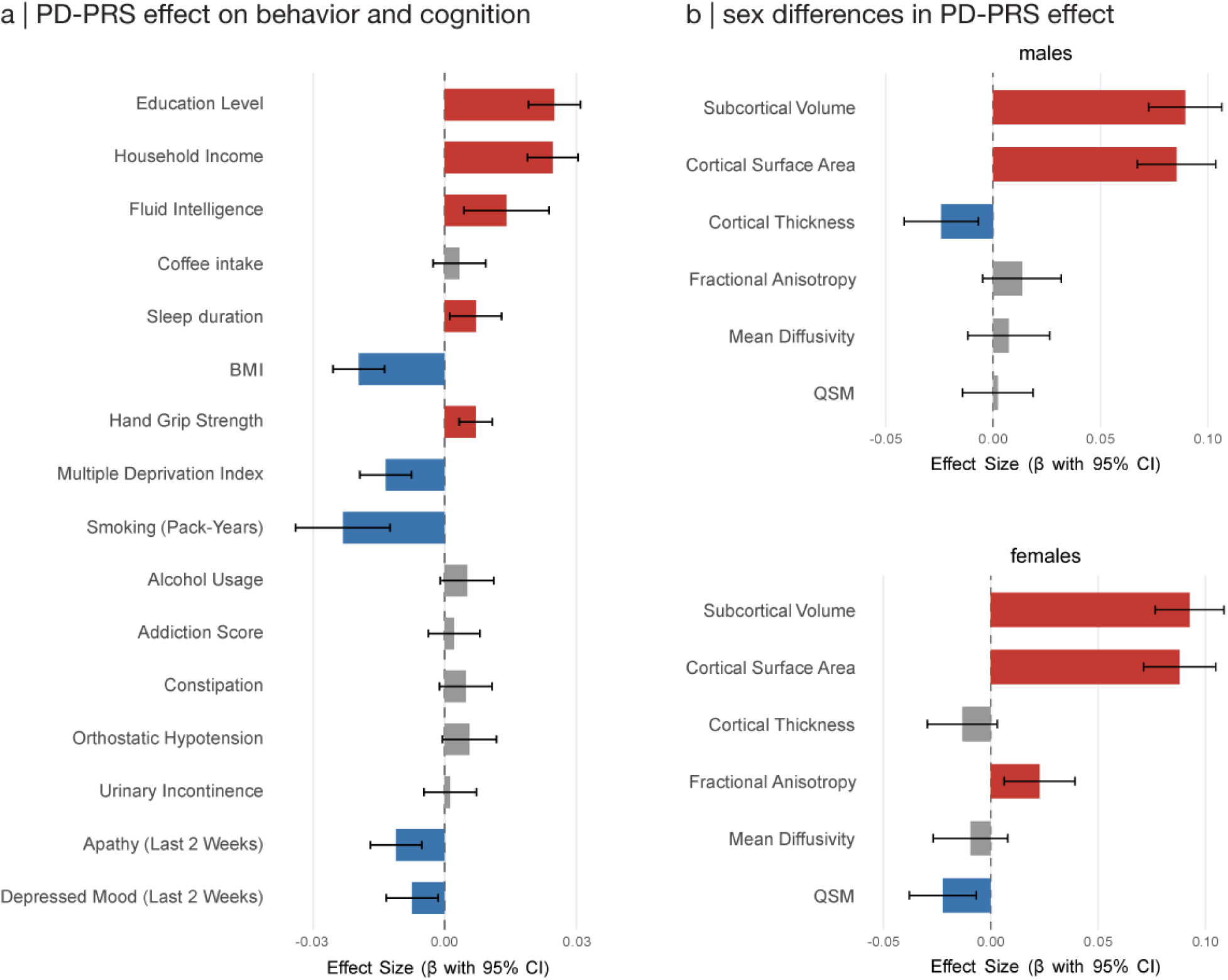
Behavioral correlates of PD-PRS and sex differences in PD-PRS brain associations. **(A)** Effect size and 95% confidence intervals showing the association between PD-PRS and characteristic PD phenotypes in healthy participants of the UK Biobanks. **(B)** Sex-specific associations between PD-PRS and global brain measures showing effect sizes and 95% confidence intervals for males and females. Only associations with pFDR<0.05 are colored. BMI: body mass index; QSM: quantitative susceptibility mapping.

### Sex-specific neuroanatomical correlates of the genetic risk for PD

PD affects males more than females ^69^ and males show a more severe course and greater brain atrophy on MRI ^70^. Therefore, we assessed whether genetic risk of PD was expressed differently across sexes. PD-PRS effects on cortical surface area and subcortical volume were comparable between males and females (Figure 3b). Cortical thickness associations were slightly more widespread in males, whereas associations with white matter FA and subcortical QSM were more pronounced in females (Supplementary Results; Supplementary Figure 3; Supplementary Table 6). These findings suggest that the sex differences in PD incidence are not driven by differential genetic effects on brain size, implicating other biological or environmental factors.

### Pathway PD-PRS associations with neuroanatomical measures

Genetic risk may render the brain vulnerable to neurodegeneration via different interacting biological mechanisms. To dissect these mechanisms, we constructed pathway-specific PD-PRSs for mitochondrial, autophagy, and lysosomal pathway genes (Mito-ALP; Supplementary Table 10), as well as a non-Mito-ALP PD-PRS comprising all remaining variants. Mito-ALP functions represent core cellular quality control mechanisms and their dysfunction is implicated in PD pathophysiology ^71,72^.

The effects of genetic risk of PD on brain size were largely driven by the non-Mito-ALP PD-PRS, which showed global and regional associations with cortical surface area and subcortical volume largely resembling those of the complete PD-PRS (Figure 4a; Supplementary Table 6). In contrast, the individual Mito-ALP-specific PD-PRSs showed more limited and selective positive associations, primarily between the mitochondrial PD-PRS and both subcortical volume and white matter FA as well as between the autophagy PD-PRS and both subcortical volume and cortical surface area. The lysosomal PD-PRS did not show any significant associations (Figure 4a; see Supplementary Results for regional results; Supplementary Table 6). To assess whether the limited associations observed for individual Mito-ALP PD-PRSs reflected reduced power due to fewer SNPs, we constructed a PD-PRS using only the top 687,574 non-Mito-ALP SNPs, matching the number of SNPs in the combined Mito-ALP PD-PRS. This PD-PRS showed global and regional associations largely similar to those of the total PD-PRS (Figure 4a; Supplementary Figure 4), suggesting that the more restricted associations seen for individual Mito-ALP pathways are not simply due to reduced SNP number. We repeated similar analyses using the PRSice method, and the results were largely consistent with those reported above (Supplementary Tables 11-12). These findings suggest that distinct PD pathways have distinct neuroanatomical signatures, with non-Mito-ALP genes predominantly driving the association between genetic risk and brain size. We therefore examined the roles of these genes and pathways in greater detail in subsequent sections.

**Figure 4.**
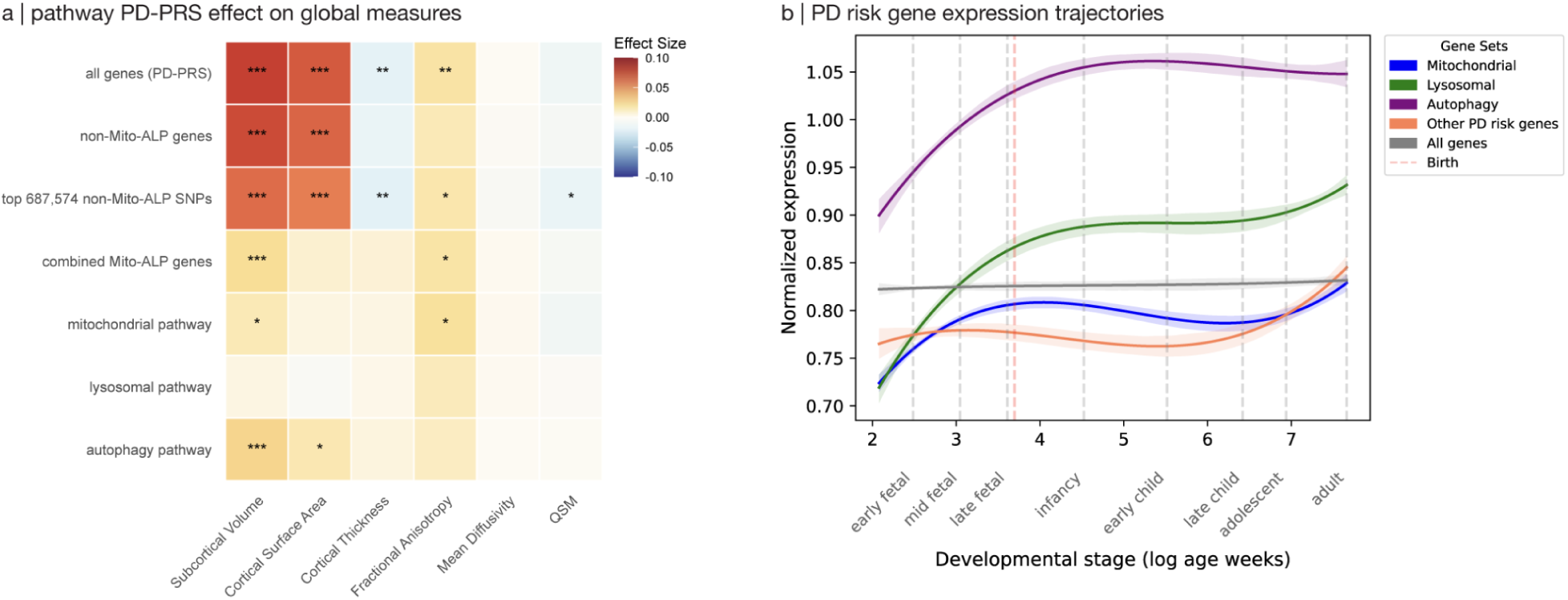
Pathway-specific PD-PRS associations and gene expression patterns. **(A)** Heatmap showing effect sizes for associations between total PD-PRS and pathway-specific PD-PRSes (rows) with global brain measures (columns). Color intensity indicates effect size with asterisks showing FDR-corrected significance levels (*p_FDR_ < 0.05, **p_FDR_ < 0.01, ***p_FDR_ < 0.001). **(B)** Gene expression trajectories across the human lifespan for pathway-specific PD risk gene sets. Lines represent the average normalized gene set expression patterns for mitochondrial, lysosomal, autophagy, and non-pathway PD risk genes from fetal stages through adulthood. The grey line represents the average expression of all genes. The red dashed line indicates birth.

### Developmental gene expression patterns of pathway-specific PD risk genes

Given the importance of Mito-ALP pathways in PD neurodegeneration and their limited associations with brain morphometry, we examined whether developmental gene expression patterns could explain the differential role of the different pathways by analyzing the expression trajectories of PD risk genes (Supplementary Tables 4 and 13) across human brain development. Our analysis revealed clear differences between Mito-ALP and non-Mito-ALP PD risk gene expression trajectories. Genes in the mitochondrial, lysosomal, and autophagy pathways had lower prenatal expression, with levels gradually increasing to reach their highest values in older age (Figure 4b). In contrast, the non-Mito-ALP genes had higher expression levels before birth (Figure 4b). The average expression trajectory across all genes further confirmed that the observed trajectories reflect pathway-specific patterns rather than a general developmental trend. These patterns suggest a dichotomy between gene sets such that while Mito-ALP genes become most active in later life, non-Mito-ALP genes are most active during the neurodevelopmental period, consistent with an influence on brain growth.

### Neuroanatomical associations of PD-PRS in ABCD developmental cohort

To test whether the neuroanatomical correlates of PD-PRS reflect neurodevelopmental processes already present during childhood and adolescence, we replicated the regression analyses in a longitudinal cohort of children and adolescents (ABCD study; four time points between ages 10–16; Supplementary Methods; Supplementary Table 14). Consistent with the findings in adults, total PD-PRS was positively associated with cortical surface area and subcortical volume at all four time points both globally (all p_FDR_ < 0.01; partial r^2^_PD-PRS_= 0.004-0.014; Figure 5a-b; Supplementary Figure 5; Supplementary Table 15) and region-wise (all p_FDR_ < 0.05). No significant associations were observed with cortical thickness, FA, or MD. As in adults, these effects were predominantly driven by the non-Mito-ALP PD-PRS, with no significant associations for the individual Mito-ALP pathways (Figure 5c-d; Supplementary Table 16). These findings suggest that the influence of PD genetic risk on brain size is detectable as early as age 10, consistent with a neurodevelopmental origin.

**Figure 5.**
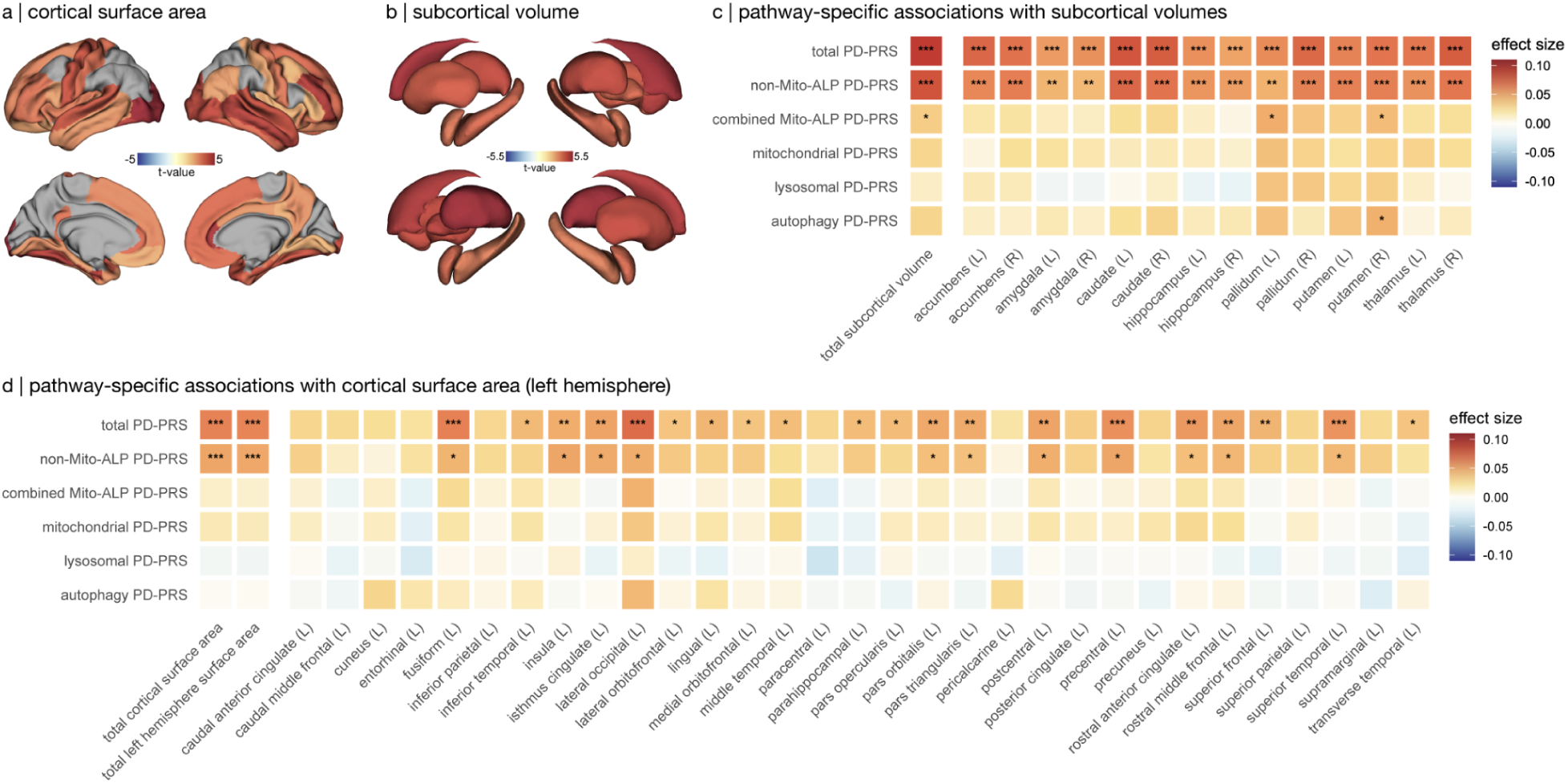
Total and pathway-specific PD-PRS associations with cortical surface area and subcortical volume at baseline visit in the ABCD dataset. **(A)** T-statistics map showing the effect of total PD-PRS on cortical surface area in the ABCD dataset. **(B)** T-statistics map showing the effect of total PD-PRS on subcortical volume in the ABCD dataset. In both maps, only regions with FDR-corrected p-value < 0.05 are colored. **(C)** Heatmap showing effect sizes for associations between total and pathway-specific PD-PRSs (rows) and global and regional subcortical volumes (columns). **(D)** Heatmap showing effect sizes for associations between total and pathway-specific PD-PRSs (rows) and global and regional cortical surface area in the left hemisphere (columns); right hemisphere results were largely consistent with those shown here. In all heatmaps, color intensity indicates effect size with asterisks denoting FDR-corrected significance levels (*p_FDR_ < 0.05, **p_FDR_ < 0.01, ***p_FDR_ < 0.001). PD-PRS: Parkinson’s disease polygenic risk score; Mito-ALP: mitochondrial, autophagy, and lysosomal pathway; ABCD: Adolescent Brain Cognitive Development.

### Pathway enrichment analysis of PD risk variants

Given the higher fetal expression of non-Mito-ALP genes, we searched for genes whose mechanisms of action linked them to specific neurodevelopmental events. Gene-set enrichment analyses using MAGMA revealed distinct biological processes for the Mito-ALP versus non-Mito-ALP PD risk variants. The Mito-ALP PD-GWAS showed enrichment primarily for neurotransmitter metabolism and oxidative stress responses, including dopamine biosynthesis, catechol-containing compound biosynthesis, tetrahydrobiopterin metabolism, and response to oxygen radicals (all p<3×10^-6^).

In contrast, the non-Mito-ALP PD-GWAS was enriched for a broader range of biological processes focused on structural cellular and developmental functions, including microtubule organization, cell division, vesicle uncoating, and endoplasmic reticulum processes (all p<2×10^-6^). Several key non-Mito-ALP genes are essential regulators of neural progenitor cell division, neuronal migration, and early synaptogenesis (e.g., TPX2, KATNB1, CLASP1, SH3GL2), and their mutations have been associated with neurodevelopmental disorders such as microcephaly and lissencephaly ^73–81^ (Supplementary Results; Supplementary Table 17). Together, these findings suggest that while Mito-ALP PD variants are mostly expressed in adulthood, non-Mito-ALP variants involve cellular processes implicated in neurodevelopment and brain growth.

### Genetic relationships between Parkinson’s disease and brain morphometry

To examine shared genetic architecture between PD risk variants and brain morphometry, we performed LDSC to estimate genetic correlations between brain imaging measures and six PD GWASs: the full PD GWAS, three individual pathway-specific GWAS subsets, a combined Mito-ALP subset, and a non-Mito-ALP subset. Both the complete and the non-Mito-ALP PD GWASs showed significant positive genetic correlations with total cortical surface area (rG > 0.226; p_FDR_ < 5.1×10^-5^) and with most individual cortical areas (0.094 ≤ rG ≤ 0.284; pFDR<0.05; Figure 6a). Similar correlation patterns were observed with subcortical volumes in all structures except the hippocampus and amygdala (0.094 ≤ rG ≤ 0.164; pFDR<0.04; Figure 6b). In contrast, the pathway-specific subsets (mitochondrial, lysosomal, autophagy, and their combination) showed very limited associations (Figure 6a-c). Genetic correlations with cortical thickness were largely non-significant across all PD GWAS subsets (Figure 6c). Together, these results indicate that PD risk variants outside the Mito-ALP pathways share genetic architecture with genes influencing brain size (surface area and subcortical volume), and that genetic influences increasing PD risk are partly shared with those promoting larger cortical surface area and subcortical volumes.

**Figure 6.**
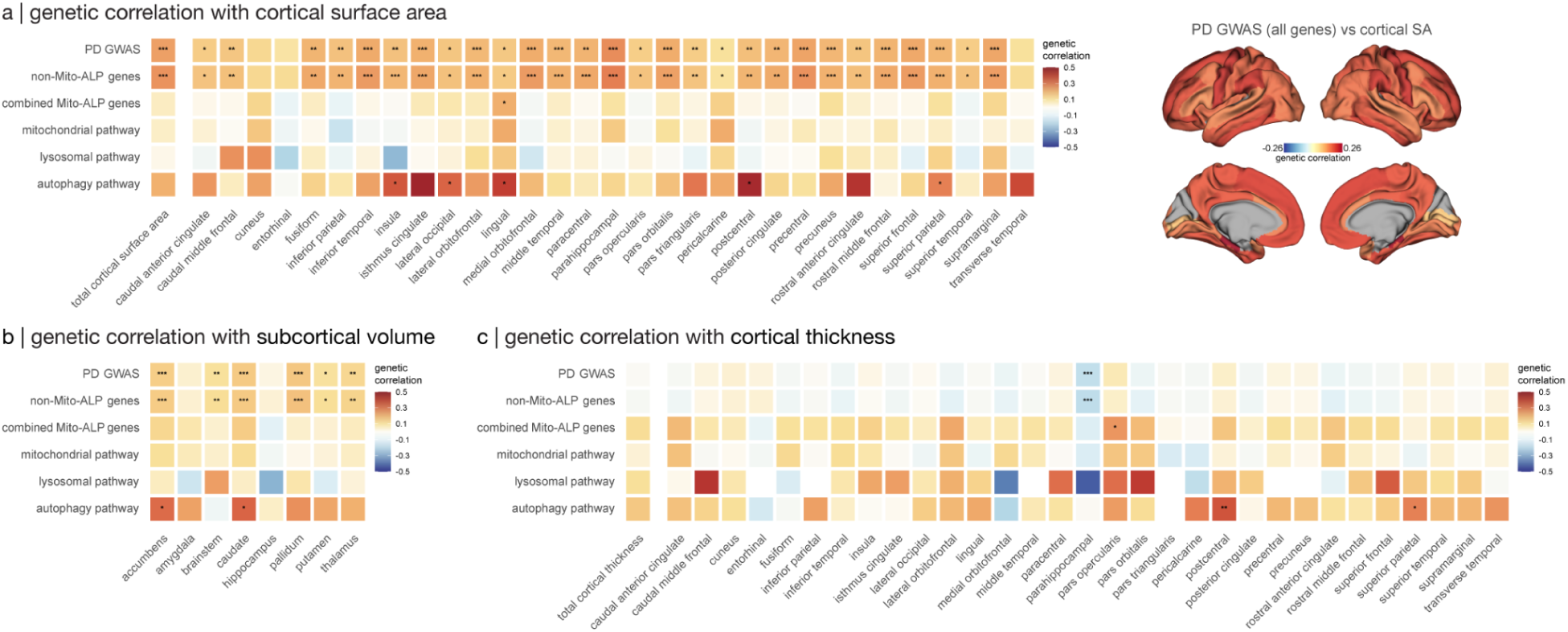
Pathway-specific PD-PRS genetic correlations with cortical and subcortical brain measures. **(A)** Left: Heatmap showing genetic correlations (rG) between total and pathway-specific PD genetic risk variants (rows) and total or regional cortical surface area (columns). Right: Brain map displaying regional genetic correlations between total PD genetic risk and cortical surface area (only genetic correlations with FDR-corrected p-value < 0.05 are colored). **(B)** Heatmap showing genetic correlations between total and pathway-specific PD genetic risk variants (rows) and regional subcortical volumes (columns). **(C)** Heatmap showing genetic correlations between total and pathway-specific PD genetic risk variants (rows) and total or regional cortical thickness (columns). In all heatmaps, color intensity represents genetic correlation strengths with asterisks denoting FDR-corrected significance levels (*p_FDR_ < 0.05, **_pFDR_ < 0.01, ***p_FDR_ < 0.001).

## Discussion

We combined genetics and neuroimaging to uncover a brain endophenotype of vulnerability to PD. Older adults at elevated genetic risk of PD did not show subclinical neuroimaging evidence of the disease. Instead, higher PD-PRS was associated with widespread increases in cortical surface area, larger subcortical volumes, and greater white matter FA. These features are not seen in PD, which is consistently associated with tissue loss. This suggests that the MRI features identified here represent a vulnerability endophenotype rather than subclinical disease. While higher genetic risk of PD was also associated with a spatially restricted pattern of lower cortical thickness in prefrontal, temporal, and limbic regions, the predominant association patterns reflect structural enlargements rather than atrophy. Consistent with evidence linking cortical surface area to cognitive ability ^82^, higher PD-PRS was also associated with higher educational attainment, greater fluid intelligence, higher household income, residing in areas with lower deprivation index, and less cigarette smoking, demographic characteristics that have also been described in PD ^83,84^. These associations suggest that higher genetic risk of PD is associated with lifelong brain and cognitive traits, possibly representing an example of antagonistic pleiotropy.

Measures of cortical surface area and thickness from T1-weighted MRI are dissociable: while both are heritable, they demonstrate little genetic overlap and are thought to result from different neurodevelopmental processes ^63,85^. This dichotomy has been linked to the radial-unit hypothesis ^86^, wherein neural progenitor differentiation in early embryogenesis determines the number of neocortical columns and hence surface area, while later developmental events influence the number of neurons and synapses per column and are reflected in cortical thickness ^66^. Furthermore, in later life, age or disease-related neurodegeneration result in reductions in cortical thickness, not surface area ^87^. A GWAS study of cortical morphometry confirmed the ontogenetic dichotomy between thickness and surface area ^63^: both anatomical features were related to genetic regulatory sites, but surface area was associated with elements active in the mid-fetal period of development while thickness was mostly linked to regulatory activity in adulthood. This is consistent with our findings that the genes from PD-PRS associated with cortical surface area were most expressed in the fetal period.

The genetically-mediated increases in brain size described here appear to directly favour later neurodegeneration. Comparisons with brain atrophy maps from ENIGMA PD revealed that areas showing the highest surface area expansion due to genetic risk of PD spatially overlapped with areas showing the most cortical thinning in PD. Thus, brain regions with greater expansion during development may be more vulnerable to damage later in life. Mendelian randomization confirmed that larger brain surface area and subcortical volumes may causally increase PD risk.

The effects of PD-PRS on brain morphology described here point toward partial neurodevelopmental origins of PD vulnerability. Replication of these associations in children aged 10–16 from the ABCD Study confirms that the influence of PD genetic risk on brain size is already present before adulthood. The global and widespread increases in cortical surface area associated with PD-PRS suggest that some PD variants exert their influence during critical early developmental windows, likely through alterations in neural progenitor cell proliferation. Higher white matter FA and subcortical volume associations with PD-PRS further support this interpretation, as a larger number of neural units entail more extensive cortico-subcortical projections. The widespread distribution of these associations across the entire brain suggests that genetic risk of PD influences fundamental neurodevelopmental processes rather than specific brain regions.

The pathway specific analyses showed that not all risk genes affect brain size. Indeed, genetic variants belonging to the mitochondrial, lysosomal, and autophagy pathways^71,72^ had limited associations with brain structure in this sample. This is consistent with gene expression trajectory analyses demonstrating that Mito-ALP PD risk genes have lower expression during fetal stages with peak expression in later life, as opposed to non-Mito-ALP PD genes which have the highest expression in the prenatal period. This distinction is further supported by gene set enrichment analyses revealing that while Mito-ALP-specific variants are enriched for neurotransmitter metabolism and oxidative stress responses, non-Mito-ALP PD risk variants primarily target neurodevelopmental processes such as cell division and microtubule organization. This dissociation suggests two distinct vulnerability mechanisms at different life stages, with non-Mito-ALP PD genes being actively involved in shaping brain structure during early development, and Mito-ALP genes primarily influencing cellular maintenance, proteostasis and metabolic processes that become important later in life when PD usually develops. Genetic correlation analyses confirmed this distinction, showing that non-Mito-ALP PD variants shared similar genetic architecture with cortical and subcortical volumes, while Mito-ALP-specific variants had minimal genetic overlap with these neuroanatomical measures. These findings support a dual vulnerability model for PD, where some genetic variants act early in development leading to larger brain structures, while others act later in life by affecting mechanisms such as protein clearance, autophagy, and mitochondrial function. However, it is important to note that the variants related to these different mechanisms are not entirely distinct, as autophagy and lysosomal pathways may also support brain development, for instance, through regulation of neural progenitor cells ^88^.

Our results have implications for epidemiological studies. For example, the often demonstrated and strong inverse association between smoking and PD is sometimes attributed to neuroprotective effects of cigarettes or to premorbid low dopamine signaling. However, another possibility raised here is the existence of genetic pleiotropy between PD and smoking propensity. Indeed, educational attainment is causally linked to reduced smoking initiation ^89^. The association between PD-PRS and neighborhood deprivation index suggests a similar type of confounding could affect associations between where people live and PD incidence.

Current evidence indicates that PD pathogenesis follows a sequential cascade of (1) initiation of alpha-synuclein misfolding, (2) self-sustaining templating, propagation and accumulation, and finally (3) neurotoxicity ^90^. Polygenic risk may affect all these processes to favour development of the disease. The autophagy, mitochondrial and lysosomal systems are implicated in mechanisms 2 and 3, and presumed to act once the pathology has been initiated; however, the portion of genetic risk mechanism related to increased brain size is presumed to exist throughout life, and it is not clear how it makes the brain vulnerable to PD. A denser neuronal architecture may favour the spread of abnormal alpha-synuclein, which is known to propagate via neuronal connections in an activity-dependent manner ^91^. Also, a larger more connected brain may be better at computation and information processing, but this may come at the cost of a precarious energetic status. Indeed, it has been shown that dopamine and noradrenaline neurons in the CNS are especially prone to oxidative stress because of their extensive arborization and number of neurotransmitter release sites, accounting for their selective vulnerability in PD ^92^. It is possible that the larger cortical and subcortical neuronal densities with higher PD PRS demonstrated here are associated with greater neurotransmission and bioenergetic demands. This may be especially relevant for dopamine neurons, whose degree of arborization is inversely associated with respiratory reserve ^93^, making densely projecting neurons vulnerable to cascading failure ^94^ after the initiation of oxidative stress and mitochondrial dysfunction. It is notable however, that the opposite argument has been made for Alzheimer’s disease, where cortical volume and educational attainment are shown to be protective, likely by raising the threshold to dementia diagnosis ^95^. The difference with PD may reflect different neuronal mechanisms of proteostatic toxicity in the two diseases. However, it is interesting to note that cognitive reserve and brain volume are also associated with faster disease progression once Alzheimer’s disease starts ^95^.

Certain limitations must be noted. The effect sizes of genetic influences on brain morphometry are small, which raises the possibility of Type I error. However, the PRS-brain associations are present in almost all cortical regions, white matter tracts and subcortical nuclei, reducing the likelihood of false positives. The replication of the genetic association with brain size in children from ABCD also strengthens the neurodevelopmental hypothesis. Another limitation is that the pathway-specific analyses are limited by the smaller number of SNPs contributing to each of the Mito-ALP PD-PRSs, which reduces statistical power and may contribute to the more limited neuroanatomical associations observed for these pathways. However, we addressed this by constructing a SNP-matched PD-PRS using non-Mito-ALP variants, which showed associations comparable to the total PD-PRS, suggesting that the restricted findings for individual pathways are not solely due to reduced SNP number. Nevertheless, the convergent evidence from gene expression trajectories, MAGMA enrichment analyses, and LDSC genetic correlations collectively support a biological distinction between these pathway-specific gene sets.

## Supporting information

Supplemental Methods and Results

## Acknowledgements

HA was supported by the Fonds de Recherche du Québec–Santé (doi.org/10.69777/366980) and Parkinson Canada. MP was supported by the Healthy Brains, Healthy Lives initiative and the Harold and Audrey Fisher Training Studentship. RM was supported by the Fonds de Recherche du Québec–Santé (https://doi.org/10.69777/350961). GS was supported by a postdoctoral fellowship from the Canadian Institutes of Health Research. NJ was supported in part by grants including RF1NS136995, R01NS107513, R01AG087513, and R01MH134004 from the National Institutes of Health. ZG was supported by grants from the Galen and Hilary Weston Foundation and the Michael J. Fox Foundation. Additionally, the G-Can (GBA1-Canada) Initiative, an open-science initiative aimed at supporting research on GBA1-associated neurodegeneration has made contributions to this research. G-Can is supported by The Hilary and Galen Weston Foundation, Silverstein Foundation, and J. Sebastian van Berkom and Ghislaine Saucier. YZ was supported by research funds from the Canadian Institutes of Health Research, Natural Sciences and Engineering Research Council of Canada, as well as Fonds de Recherche du Québec-Santé (https://doi.org/10.69777/320107). AD was supported by research funds from the Canadian Institutes of Health Research, the Michael J Fox Foundation for Parkinson’s Research and a Canadian Institutes of Health Research Foundation Award.

## Author Contributions

Conceptualization: HA, NA, YZ, AD

Data curation: HA, NA, YZ, AD

Formal analysis: HA, NA, LL, APB, CT

Funding acquisition: YZ, AD

Investigation: HA, NA, APB, CT

Methodology: HA, NA, LL, APB, CT, MP, KS, FM, AV, RM, EY, PS, RDM, GS, NK

Project administration: HA, NA, YZ, AD

Resources: HA, NA, LL, RM, JBP, YZ, AD

Software: HA, NA, LL, APB, CT, MP, FM, RM, PS, RDM

Supervision: YZ, AD

Validation: HA, NA

Visualization: HA, NA

Writing – original draft: HA, NA, LL, YZ, AD

Writing – review & editing: HA, NA, KS, FM, AV, NJ, PMT, ZGO, BM, YZ, AD

## Data Availability

All data are linked to in the main text or are available in the supplementary materials. All codes are available on GitHub (https://github.com/HoumanAzizi).

