## Supplemental Methods and Results for "Brain size links neurodevelopment to neurodegeneration in Parkinson’s disease"

### Supplementary Methods

*Detailed exclusion criteria*

The following exclusion criteria were applied: history of bipolar or any neurological disorder, including Parkinson's disease, dementia and cognitive impairment, Alzheimer's disease, chronic/degenerative neurological problem, acute infective polyneuritis or Guillain-Barré syndrome, multiple sclerosis, other demyelinating disease (not multiple sclerosis), stroke, brain hemorrhage, brain abscess/intracranial abscess, cerebral aneurysm, cerebral palsy, encephalitis, epilepsy, head injury, infection of the nervous system, ischemic stroke, meningioma/benign meningeal tumor, meningitis, motor neuron disease, neurological injury/trauma, spina bifida, subdural hemorrhage/hematoma, transient ischemic attack (TIA), subarachnoid hemorrhage (UKB field ID: 20002), and bipolar disorder type I and II (UKB field ID: 20126); first-degree family history of Parkinson's disease (UKB field IDs: 20107, 20110, 20111); BMI > 35 (UKB field ID: 21001); relation to another participant closer than cousin (UKB field ID: 22011); genetic and self-reported sex mismatch (UKB field IDs: 22001, 31); and non-European genetic ethnicity (UKB field ID: 21006). The European-ancestry restriction was applied to ensure validity of genetic analyses as the input GWAS were performed in European-ancestry individuals. In addition, participants with brain measures outside of 3 standard deviations in each modality were excluded.

*Brain imaging acquisition and preprocessing*

All participants underwent 3D volumetric brain MRI scanning on a 3T Siemens Skyra (VD13A SP4) scanner equipped with a standard Siemens 32-channel head coil, located in one of three UK sites, as detailed previously [1, 2]. Structural MRI data were acquired as high-resolution

T1-weighted images using a 3D MPRAGE sequence at 1-mm isotropic resolution with identical acquisition protocols. Data were submitted to automated preprocessing and quality control pipelines by the UKB Team [2].

Cortical thickness and surface area values were generated in-house using FreeSurfer v7.4.1 [3], with parcellation of the surface using the DKT atlas [4, 5]. Subcortical volume values were generated using FreeSurfer's aseg pipeline [6] based on the Harvard-Oxford subcortical structural atlas parcellation [7]. All FreeSurfer outputs were visually inspected and quality controlled. Data from 46,349 participants of the UKB data release in early 2021 were downloaded. After applying the exclusion criteria and quality control, we included data from 25,256 individuals for surface area, 25,237 for cortical thickness, and 25,255 for subcortical volume analysis.

3D diffusion MRI (dMRI) scans were corrected for eddy currents, head motion and slice outliers using the Eddy tool (http://fsl.fmrib.ox.ac.uk/fsl/fslwiki/EDDY) [8]. Gradient distortion correction was then applied. Quality control was performed based on the total number of outlier slices, reflective of head motion during dMRI scanning. The diffusion MRI sequence included 105 volumes acquired at 2 $mm^3$ resolution, with 5 at $b = 0$ s/$mm^2$, 50 at $b = 1000$ s/$mm^2$, and 50 at $b = 2000$ s/$mm^2$.

dMRI and T1-weighted MRI scans were processed with TractoFlow v2.2.1 [9] using default parameters (https://github.com/neurodatascience/tractoflow_UKBB) to obtain a white matter tractogram and standard diffusion maps for FA and MD. White matter tractograms were used to calculate whole-brain diffusion measure maps (FA and MD) for each participant.

800 fiber clusters from the O'Donnell Research Group (ORG) anatomically curated atlas [10] were organized into 73 anatomical tracts, including 58 deep white matter tracts, major long-range association and projection tracts, commissural tracts, and tracts related to the brainstem and

cerebellar connections. All subject-specific white matter diffusion maps and the white matter anatomical tract atlases were registered to the ICBM 152 nonlinear 2009c atlas [11]. Subsequently, we obtained average diffusion measures (FA and MD) across each anatomical tract for each individual (N ≥ 23,245).

QSM MRI data for subcortical nuclei from the Harvard-Oxford subcortical atlas, as well as the substantia nigra (total of 16 nuclei) [12], were obtained directly from UKB for 30,225 participants.

*Genetic data quality control*

We considered the 487,410 samples included in the 2019 release of UKB. Genetic data were phased and imputed to ~96 million variants with the Haplotype Reference Consortium (HRC) haplotype resource and the UK10K + 1KG reference panel [13]. We restricted our sample to participants with European ancestry based on self-report and genetic principal component thresholds. We excluded individuals with relatedness closer than cousins, creating a maximally unrelated study sample. Finally, we excluded first-degree relatives of people with PD, individuals whose self-reported sex did not match genotyping, and participants with extreme heterozygosity or high genotyping missingness.

For variant-level quality control, we removed SNPs with minor allele frequency (MAF) < 1%, missing rate > 1%, imputation quality INFO score < 0.3, or significant deviation from Hardy-Weinberg equilibrium ($P < 1 \times 10^{-10}$). All genetic quality controls were conducted using PLINK 1.9 [14, 15].

the “auto” global shrinkage parameter

*PRS-CS calculation in UKB Biobank dataset*

PRS-CS [16] and PLINK 2.0 [14, 15] were used to calculate polygenic risk scores. PRS-CS uses a Bayesian regression framework with continuous shrinkage (CS) priors on SNP effect sizes. To calculate PD-PRS, posterior SNP weights were inferred by PRS-CS using the effect sizes of variants from the latest PD GWAS summary statistics [17] using "auto" global shrinkage parameter. The 1000 Genomes Project European sample served as the external linkage disequilibrium (LD) reference panel [18]. The individual-level polygenic score was generated by concatenating posterior SNP weights from all chromosomes and applying PLINK's --score command. The same approach was used for pathway-specific PD-PRS calculations, using posterior weights from the previous step for the subset of SNPs assigned to each gene set.

*PRS-ice calculation in UKB Biobank dataset*

Total and pathway-specific PRS for PD were computed using the PRSet module implemented in PRSice-2 [19]. Prior to scoring, SNPs were subjected to linkage disequilibrium (LD)-based clumping ($r^2 < 0.1$ within a 250 kb window) using the target sample as the LD reference panel, and a p-value inclusion threshold of $P \leq 0.05$ was applied to the base GWAS summary statistics [20]. PD-PRS were then calculated for each participant as the weighted sum of allele dosages, with GWAS effect sizes (log odds ratios) used as weights. All scores were standardised prior to association analyses. Pathway genes were defined as previously discussed and pathway-specific PD-PRS was calculated using the subset of SNPs assigned to each gene set.

PRS-CS calculation in ABCD dataset

We used genetic data from 11,670 participants included in the Adolescent Brain Cognitive Development (ABCD) Study. Saliva and blood samples were collected at the baseline visit and genotyped using the Affymetrix Smokescreen genotyping array (~515,000 SNPs) [21]. After dish quality control and profile checks, genotypes were called using Axiom Analysis Suite, followed by study-level quality control filtering for SNP-level missingness (<10%) and sample-level missingness (<20%) [22]. We used genetic principal components with HapMap3 data as the reference panel to identify participants of European ancestry. We excluded individuals with relatedness closer than cousins. Pre-imputation steps followed TOPMed documentation. The curated genotype data were then phased with Eagle and imputed to ~260 million variants using the TOPMed r3 reference panel on the TOPMed Imputation Server [23]. Following imputation, we applied additional variant-level quality control by removing SNPs with minor allele frequency (MAF) < 1%, missing rate > 1%, imputation quality INFO score < 0.3, or significant deviation from Hardy-Weinberg equilibrium (HWE) with $P < 1 \times 10^{-10}$.

Adolescent Brain Cognitive Development (ABCD) cohort analysis

*Participant characteristics.* To examine whether the neuroanatomical correlates of PD genetic risk seen in adults are also detectable during childhood and adolescence, we replicated the PD-PRS regression analyses in the Adolescent Brain Cognitive Development (ABCD) cohort [24–26], a prospective longitudinal, multi-site study of children recruited at ages 9–10 years across the United States. Study procedures were approved by review boards of all participating sites and written parental informed consent and child assent were collected prior to participation. We used data from four assessment time points and included only participants who had neuroimaging data that passed MRI quality control checks and had resting-state fMRI motion data available. We further restricted our participants to those of European genetic ancestry as PD GWAS was conducted in this population. To account for the family-based design of the ABCD Study, one participant was randomly retained per family unit (based on the genetic family identifier) at each time point. The final sample sizes ranged from 1,906 to 4,387 depending on the time point and modality (Supplementary Table 13), including baseline visit (age = 10.0±0.6 years), two-year follow-up (age = 12.0±0.6 years), four-year follow-up (age = 14.2±0.7 years), and six-year follow-up (age = 16.1±0.6 years).

*Brain imaging acquisition and preprocessing.* All participants were scanned on 3T MRI scanners (Siemens, General Electric, and Philips) at 22 sites using a harmonized acquisition protocol [24]. We used cortical surface area, cortical thickness, subcortical volume, and white matter fractional anisotropy (FA) and mean diffusivity (MD) data provided by the ABCD initiative [27]. Cortical surface area and thickness values were parcellated using the Desikan-Killiany-Tourville (DKT) atlas (68 cortical regions) [5], and subcortical volumes were derived using FreeSurfer's automated subcortical segmentation [6]. Diffusion MRI data were processed to yield fractional anisotropy (FA) and mean diffusivity (MD) measures averaged across white matter tracts [28]. Data were harmonized using ComBat harmonization software to remove site variability from each brain measure [29].

*Spatial correlation analyses*

We compared spatial patterns of PD-PRS cortical associations against patterns of cortical atrophy observed in PD, using cortical thickness and surface area maps from the ENIGMA-PD consortium reflecting alterations in 3,096 people with PD compared to healthy controls at each Hoehn and Yahr disease stage, while accounting for age and sex (34). To account for spatial autocorrelation among brain regions [30], we evaluated statistical significance using spatial null models (spin-test) [31]. Null models were generated using the netneurotools toolbox (https://netneurotools.readthedocs.io/en/latest/). DKT-atlas cortical surface parcel coordinates [4] were projected to the surface of spheres and randomly rotated 10,000 times. Cortical surface data were then reassigned with the values of the closest rotated parcel, constructing a null distribution that preserved the spatial autocorrelation of the original surface map and providing a null model benchmark against which we compared the observed spatial correlations.

*Behavioral trait definitions*

We investigated the association between PD-PRS and the following behavioral phenotypes in the neurologically and psychologically healthy participant from the entire UKB dataset: education level (UKB field ID: 6138; Number of years of education defined based on the International Standard Classification of Education (ISCED) scale [32] as: college or university degree = 20 years; NVQ or HND or HNC or equivalent = 19 years; other professional qualifications, e.g., nursing, teaching = 15 years; A levels/AS levels or equivalent = 13 years; O levels/GCSEs or equivalent = 10 years; CSEs or equivalent = 10 years; none of the above = 7 years), household income (UKB field ID: 738, defined by taking median of each category as: Less than 18,000 = 9,000; 18,000 to 30,999 = 24,500; 31,000 to 51,999 = 41,500; 52,000 to 100,000 = 76,000; Greater than 100,000 = 125,000; in pounds), fluid intelligence (UKB field ID: 20016; the capacity to think logically and solve problems in novel situations, independent of acquired knowledge), coffee intake (UKB field ID: 1498; number of cups per day), sleep duration (UKB field ID: 1160; average hours of sleep per day), measured BMI (UKB field ID: 21001, in $kg/m^2$), multiple deprivation index (UKB field ID: 26410; a composite measure generated by the UK government reflecting the level of deprivation associated with each small geographic area in the UK), pack years of cigarette smoking (UKB field ID: 20161), alcohol usage (UKB field IDs: 20414, 20403; calculated through multiplying frequency of drinking alcohol by amount of alcohol drunk on a typical drinking day; in units of alcohol per month), addiction score (UKB category: 141; calculated as the total number of behavioral, medication, and substance addictions), hand grip strength (UKB field IDs: 46, 47; calculated as the average strength in both hands), constipation, orthostatic hypotension, urinary incontinence (UKB field ID: 41202; based on the International Classification of Diseases version-10 (ICD-10) diagnosis codes K59.0, I95.1, and N39.3, N39.4, and R32), and apathy and depressed mood (UKB field IDs: 2060, 2050; frequency in last 2 weeks defined as: not at all = 0; several days = 5; more than half the days = 10; nearly every day = 14).

*Mendelian randomization parameters*

Two-sample Mendelian randomization (MR) is a statistical method that uses genetic variation as an instrumental variable to investigate the causal influence of an exposure variable on an outcome. Here, two-sample MR was used to discover whole-brain and region-wise neuroanatomical measures that may have a causal effect on PD diagnosis.

All MR analyses were performed using the TwoSampleMR package in R [33, 34]. For each exposure, significant SNPs ($p < 5 \times 10^{-8}$) were selected as genetic instrumental variables. The following values were extracted: rs-numbers, beta coefficients, standard errors, p-values, sample sizes, allele frequencies, and effect alleles. SNPs within 10,000 kb of each other or with $R^2 > 0.001$ were clumped to account for linkage disequilibrium (LD). Average $R^2$ values were then calculated for each set of instrumental SNPs, reflecting the average proportion of variance in exposure explained by the selected SNPs [35]. Instrumental SNPs were harmonized between exposure and outcome GWASs, and Steiger filtering was applied to exclude SNPs explaining more variance in the outcome than in the exposure.

Inverse-variance weighted (IVW), MR Egger, and weighted median estimators were performed on each exposure–outcome pair to account for heterogeneity and horizontal pleiotropy [36–39]. Heterogeneity and pleiotropy were assessed to ensure the validity of causal estimates, as heterogeneity may indicate instrument strength variations, while horizontal pleiotropy violates the MR assumption that genetic variants affect the outcome only through the exposure variable.

Reverse MR was performed to test whether PD diagnosis has a causal effect on brain structure, using PD as the exposure and each whole-brain neuroanatomical measure as the outcome. Instrumental SNPs were selected from the PD GWAS at genome-wide significance of $p < 5*10^{-8}$ and clumped for LD as described above. SNPs were harmonized between the PD exposure and neuroimaging outcome GWASs, and the same set of above-mentioned MR methods were applied to assess potential reverse causation.

*BrainSpan gene expression normalization and calculation*

To create gene expression trajectories, we used the open-access BrainSpan RNA-sequencing dataset (Gencode v10 summarized to genes), containing developmental transcriptomics of the human brain from pre-natal stages to adulthood [40]. The data included 524 samples from 26 cortical and subcortical regions from 42 donors across 31 developmental stages spanning 8 weeks post-conception (PCW) to 40 years of age. The RNA-sequencing matrix contained normalized RPKM (Reads Per Kilo-base per Million) values for 52,376 genes across 524 samples. These values were normalized using conditional quantile normalization to account for GC content and sequencing depth and batch effect correction using ComBat [41, 42].

We performed cleanup of the sample-by-gene expression matrix using only cortical samples: (1) samples were grouped into major developmental stages including early fetal, mid fetal, late fetal, infancy, early childhood, late childhood, adolescence, and adulthood [43]; (2) regions with at least 1 sample in each age group were retained; (3) duplicate genes were removed, yielding 47,808 unique genes; (4) genes were retained if they had RPKM $\geq 1$ in 80% of samples at each spatiotemporal point [44]. This Cleanup resulted in a sample of 352 specimens (155 females) from 11 cortical regions and 8,370 genes. Cortical regions included: anterior (rostral) cingulate (medial prefrontal) cortex, dorsolateral prefrontal cortex, inferolateral temporal cortex (area TEv, area 20), orbital frontal cortex, posterior (caudal) superior temporal cortex (area 22c), posteroventral (inferior) parietal cortex, primary auditory cortex (core), primary motor cortex (area M1, area 4), primary somatosensory cortex (area S1, areas 3, 1, 2), primary visual cortex (striate cortex, area V1/17), and ventrolateral prefrontal cortex. Expression values were log2-transformed and normalized using the upper-quartile method, with each donor's data scaled by their 75th percentile expression value [45–47].

Sample-by-gene expression matrices were then retrieved for each pathway. For each sample, the mean expression across genes within each pathway was calculated, and the average pathway expression across all genes per age group was computed. Pathway expressions were then plotted across age categories, with smoothed curves were produced using a cubic polynomial fit.

*MAGMA parameters*

We separated the latest PD GWAS summary statistics [17] into two subsets: a Mito-ALP-specific PD GWAS containing only SNPs mapped to lysosomal, mitochondrial, and autophagy pathway genes, and a non-Mito-ALP PD GWAS containing all remaining SNPs. The distinct biological signatures of each subset were investigated using separate MAGMA analyses.

All SNPs were mapped to protein-coding genes (Ensembl reference gene v102) with an extended window of 35 kb upstream and 10 kb downstream of each gene to capture regulatory regions. Gene-level association statistics were calculated using MAGMA's default SNP-wise mean model with default parameters. Competitive gene-set analysis was subsequently performed. We evaluated over 15,000 gene sets from the Molecular Signatures Database (MSigDB v2023.1.Hs), including curated gene sets from KEGG, Reactome, and BioCarta, and Gene Ontology (GO) terms across biological processes, cellular components, and molecular functions categories [48, 49]. Statistical significance was assessed using Bonferroni correction across the number of gene sets tested, with Bonferroni-corrected $p < 0.05$ being considered as significant.

### Supplementary Results

*Sex-specific differences in neuroanatomical correlates of the genetic risk for PD*

The PD-PRS effects on cortical surface area were similar between males and females (r=0.71; $p_{spin}$=0.0003), with comparable global effect sizes (Figure 4b) and regional patterns (Supplementary Figure 3; Supplementary Table 6) for both cortical surface area and subcortical volumes. Considering cortical thickness, PD-PRS was negatively associated with several cortical regions in both sexes, although associations were slightly more widespread and pronounced in males (r=0.49; $p_{spin}$=0.0011; Supplementary Figure 3). In contrast, PD-PRS showed more pronounced associations with white matter FA and subcortical QSM in females, with significant global effects (Figure 4b) and more widespread regional associations present in females but not in males (Supplementary Figure 3; Supplementary Table 6).

*Regional pathway-specific PD-PRS associations with brain morphometry*

At the regional level, the mitochondrial PD-PRS was positively associated with FA across multiple tracts while the autophagy PD-PRS was positively associated with subcortical volumes across multiple structures (all pFDR<0.05; Supplementary Table 6). The lysosomal PD-PRS did not show significant regional associations with any neuroanatomical measure.

*Neurodevelopmental roles of top non-Mito-ALP PD risk genes*

Several key genes within this pathway play critical roles in early brain development (Supplementary Table 16). *TPX2* and *KATNB1* both serve as essential regulators of neural progenitor cell divisions, with *TPX2* functioning as a microtubule nucleation factor for spindle assembly [50, 51] and *KATNB1* encoding the regulatory subunit of the microtubule-severing enzyme katanin [52, 53]. *CLASP1* drives microtubule dynamics during neural differentiation and has been implicated in axon guidance and neuronal migration [54]. Mutations in these genes have further been shown to influence brain development leading to neurodevelopmental delay and disorders such as microcephaly and lissencephaly [55]. Another top hit is *SH3GL2* (endophilin A1) which mediates synaptic vesicle endocytosis and is required for normal synaptogenesis, dendrite outgrowth, and neural circuit formation during early synaptic development [56–58].

### Supplementary Figures

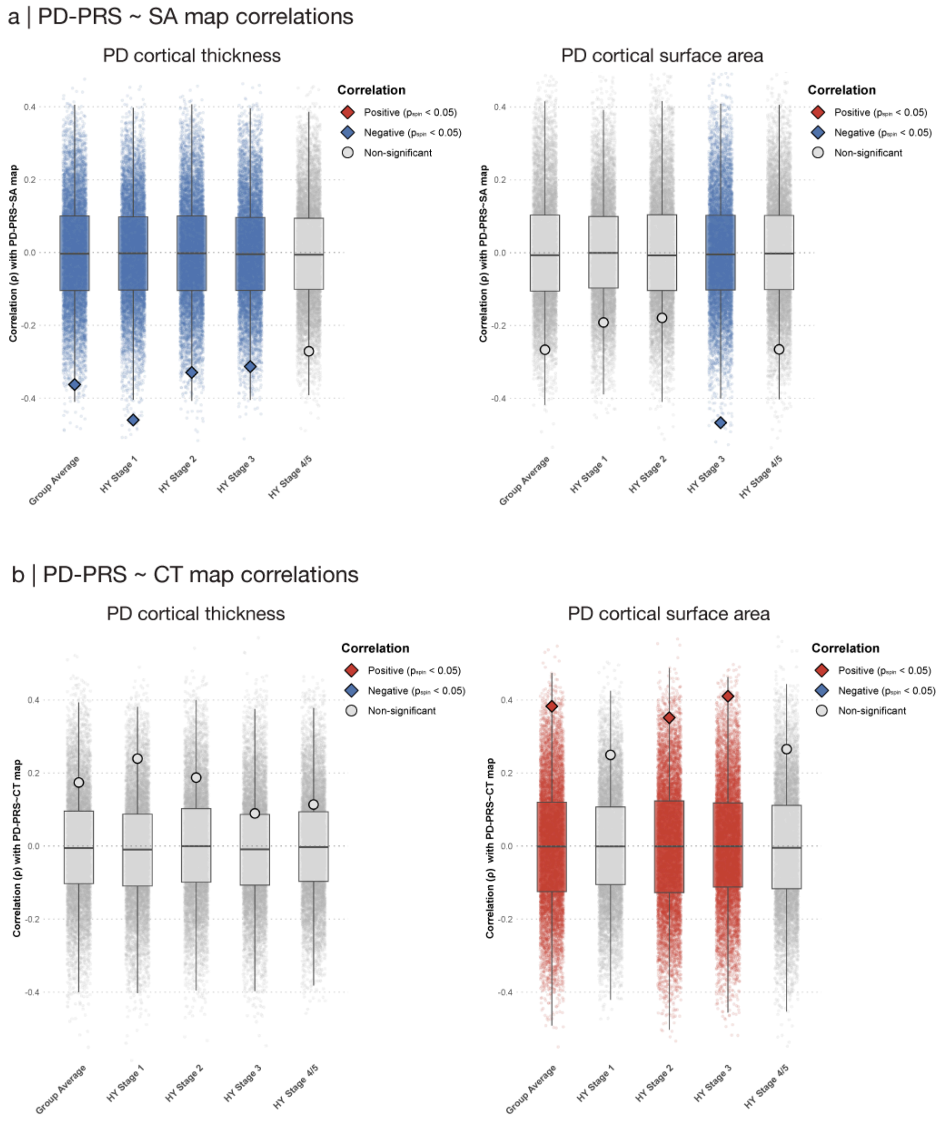


**Supplementary Figure 1. Comparison of the spatial distribution of the PD-PRS effect on cortical surface area and thickness with atrophy patterns in Parkinson's disease.** Correlation of PD-PRS effect on cortical surface area **(A)** and cortical thickness map **(B)** with PD cortical thickness (left column) and surface area maps (right column) from ENIGMA (PD minus controls) across HY disease stages and group average. Box plots show the spatial null distributions created using spin-test, with larger squares (significant) and circles (non-significant) showing the actual correlations between corresponding maps. HY: Hoehn & Yahr disease stages.

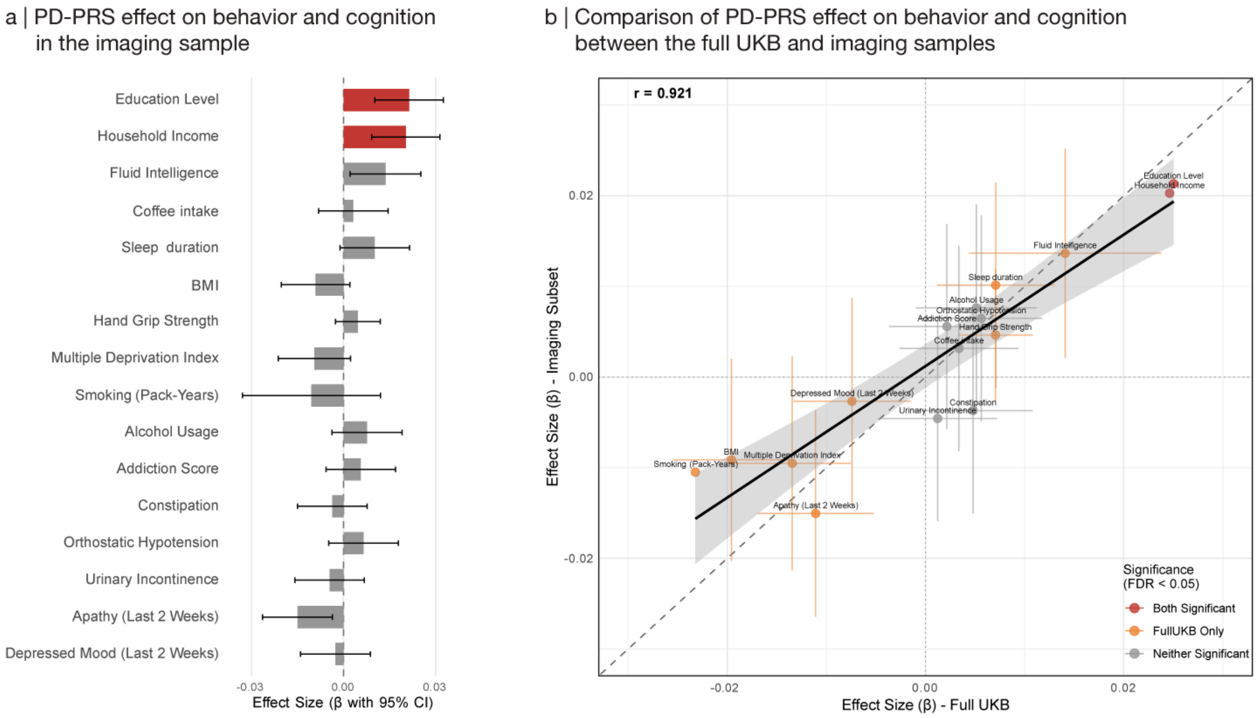


**Supplementary Figure 2. Behavioral correlates of PD-PRS in the imaging sample. (A)** Effect size and 95% confidence intervals showing the association between PD-PRS and characteristic PD phenotypes in healthy imaging participants of the UK Biobanks. **(B)** Comparison of the effect sizes of the association between PD-PRS and characteristic PD phenotypes between the full healthy UK Biobank (x-axis) and imaging (y-axis) samples. BMI: body mass index.

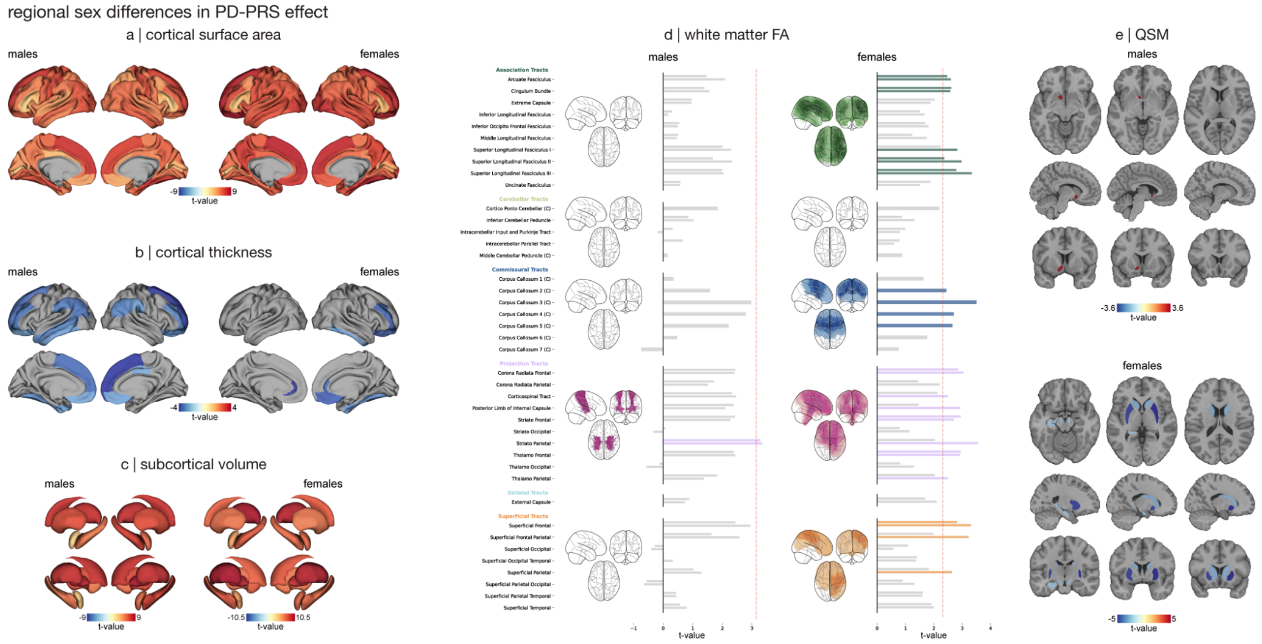


**Supplementary Figure 3. Sex-specific association between PD-PRS and morphometrical brain measures. (A-C)** T-statistics maps showing the effect of PD-PRS on cortical surface area, cortical thickness, and subcortical volume in males (left) and females (right). **(D)** Glass brain and bar plots showing t-statistics for the effect of PD-PRS on white matter tract FA in males (left) and females (right). Each glass brain shows projection of significant tracts in its corresponding tract category. Bar plots are shown as a single bar for cross-hemispheric tracts and as two bars for bilateral tracts (left hemisphere: upper bar; right hemisphere: lower bar). The red dashed line shows the FDR significance threshold. **(E)** T-statistics maps showing the effect of PD-PRS on subcortical QSM in males (top) and females (bottom). In all plots, only regions with FDR-corrected p-value < 0.05 are colored. PD-PRS: Parkinson's disease polygenic risk score.

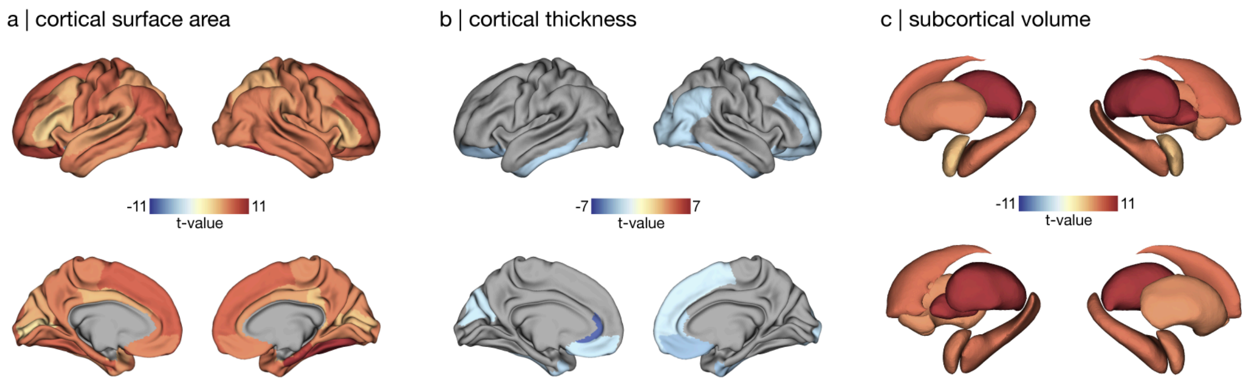


**Supplementary Figure 4. Association between SNP-count-matched PD-PRS regionwise neuroanatomical measures in the UK Biobank.** T-statistics maps showing the effect of pathway PD-PRS using the top 687,574 PD SNPs on **(A)** cortical surface area, **(B)** cortical thickness, and **(C)** subcortical volume. In all plots, only regions with FDR-corrected p-value < 0.05 are colored. PD-PRS: Parkinson's disease polygenic risk score.

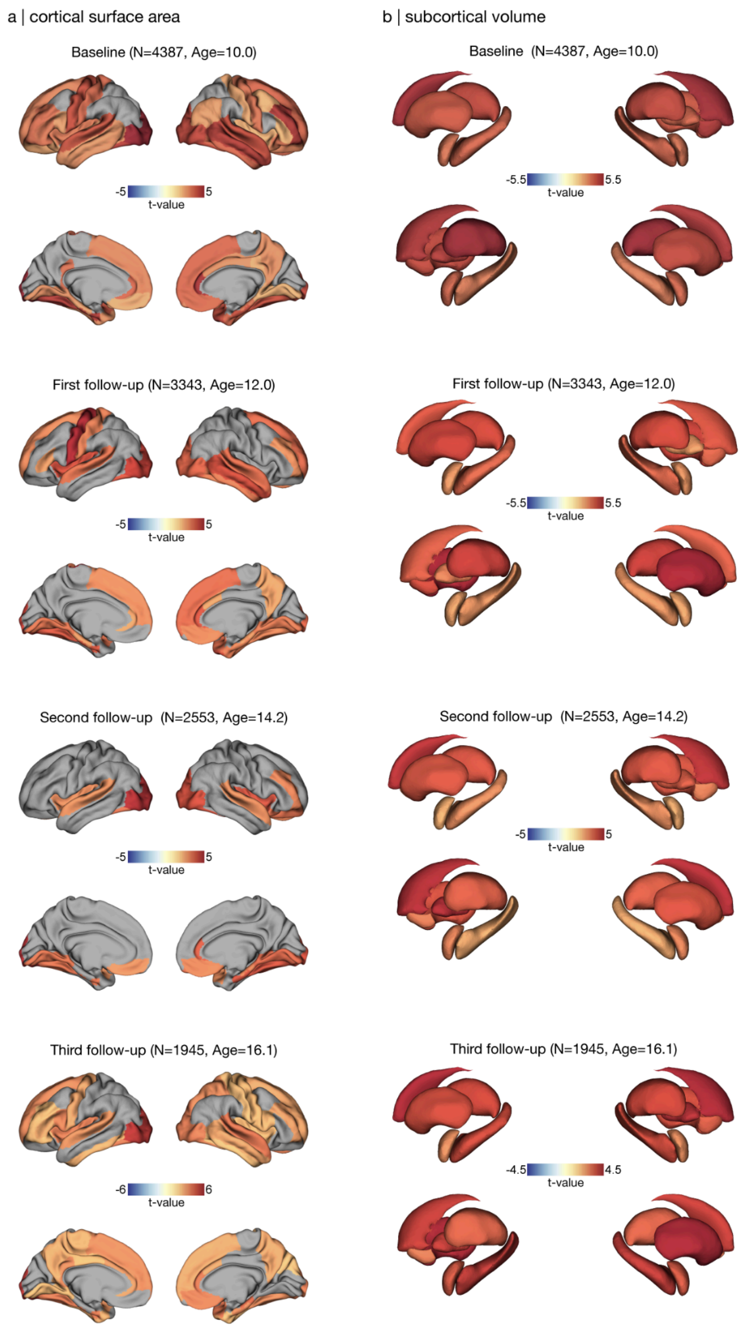


**Supplementary Figure 5. Association between total PD-PRS and cortical surface area and subcortical volume across all time points in the ABCD dataset.** T-statistics maps showing the effect of total PD-PRS on **(A)** cortical surface area and **(B)** subcortical volume at baseline and first, second, and third follow-up visits (rows). In all maps, only regions with FDR-corrected p-value $< 0.05$ are colored. PD-PRS: Parkinson's disease polygenic risk score; ABCD: Adolescent Brain Cognitive Development.

**Supplementary Tables**

**Supplementary Table 1.** Demographics and number of UK Biobank participants excluded or remaining in analyses after applying each exclusion criteria.

**Supplementary Table 2.** Total number of SNPs per pathway in mitochondrial, lysosomal, autophagy, the combined Mito-ALP pathway, the non-Mito-ALP pathway, as well as the complete PD GWAS.

**Supplementary Table 3.** Associations between PD-PRS and behavioral and demographic characteristics in the full and neuroimaging subset of UK Biobank participants.

**Supplementary Table 4.** Classification of Parkinson's disease risk genes into mitochondrial, lysosomal, autophagy, and non-Mito-ALP pathway categories.

**Supplementary Table 5.** Final sample sizes for each analysis across imaging modalities and cohorts.

**Supplementary Table 6.** Statistical results of the associations between total and pathway-specific PD-PRS and all regional brain measures including cortical thickness, surface area, subcortical volumes, white matter diffusion metrics, and quantitative susceptibility mapping values across males, females, and both sexes combined in the UK Biobank dataset.

**Supplementary Table 7.** Variance explained for global brain measures by regression models and the partial variance explained by PD-PRS after adjusting for covariates.

**Supplementary Table 8.** Spatial correlation results between PD-PRS cortical thickness and surface area effect maps and ENIGMA-PD atrophy maps across Hoehn and Yahr disease stages.

**Supplementary Table 9.** Mendelian randomization results including instrumental variable statistics, heterogeneity tests, pleiotropy assessments, and causal estimates for associations between brain structural measures and PD risk.

**Supplementary Table 10.** List of genes in mitochondrial, lysosomal, and autophagy pathways.

**Supplementary Table 11.** Statistical results of the associations between total and pathway-specific PD-PRS and all global brain measures using the PRSice-2 method.

**Supplementary Table 12.** Statistical results of the associations between total and pathway-specific PD-PRS and all regional brain measures using the PRSice-2 method.

**Supplementary Table 13.** Number of genes in mitochondrial, lysosomal, autophagy, and the combined Mito-ALP pathways and the subset identified as PD risk genes in each pathway.

**Supplementary Table 14.** Sample characteristics and demographics of participants from the Adolescent Brain Cognitive Development (ABCD) study included in longitudinal neuroimaging analyses.

**Supplementary Table 15.** Statistical results of the associations between total and pathway-specific PD-PRSs and global brain measures across four longitudinal time points in the ABCD cohort.

**Supplementary Table 16.** Statistical results of the associations between total and pathway-specific PD-PRSs and regional brain measures across four longitudinal time points in the ABCD cohort.

**Supplementary Table 17.** Key neurodevelopmental genes identified in pathway enrichment analysis of non-Mito-ALP PD risk variants.

27. Hagler DJ Jr, Hatton S, Cornejo MD, Makowski C, Fair DA, Dick AS, et al. Image processing and analysis methods for the Adolescent Brain Cognitive Development Study.

Neuroimage. 2019;202:116091.

36. Bowden J, Davey Smith G, Burgess S. Mendelian randomization with invalid instruments: effect estimation and bias detection through Egger regression. Int J Epidemiol.

2015;44:512–525.

37. Bowden J, Davey Smith G, Haycock PC, Burgess S. Consistent estimation in Mendelian randomization with some invalid instruments using a weighted median estimator. Genet Epidemiol. 2016;40:304–314.

38. Burgess S, Bowden J, Fall T, Ingelsson E, Thompson SG. Sensitivity analyses for robust causal inference from Mendelian randomization analyses with multiple genetic variants. Epidemiology. 2017;28:30–42.

39. Burgess S, Butterworth A, Thompson SG. Mendelian randomization analysis with multiple genetic variants using summarized data: Mendelian randomization using summarized data. Genet Epidemiol. 2013;37:658–665.

40. Allen Institute for Brain Science. Allen Developing Human Brain Atlas: Developmental Transcriptome. 2010.

41. Hansen KD, Irizarry RA, Wu Z. Removing technical variability in RNA-seq data using conditional quantile normalization. Biostatistics. 2012;13:204–216.

42. Li M, Santpere G, Imamura Kawasawa Y, Evgrafov OV, Gulden FO, Pochareddy S, et al. Integrative functional genomic analysis of human brain development and neuropsychiatric risks. Science. 2018;362:eaat7615.

43. Kang HJ, Kawasawa YI, Cheng F, Zhu Y, Xu X, Li M, et al. Spatio-temporal transcriptome of the human brain. Nature. 2011;478:483–489.

44. Parikshak NN, Luo R, Zhang A, Won H, Lowe JK, Chandran V, et al. Integrative functional genomic analyses implicate specific molecular pathways and circuits in autism. Cell. 2013;155:1008–1021.

45. Bullard JH, Purdom E, Hansen KD, Dudoit S. Evaluation of statistical methods for

normalization and differential expression in mRNA-Seq experiments. BMC Bioinformatics. 2010;11:94.

52. Dunleavy JEM, O’Connor AE, Okuda H, Merriner DJ, O’Bryan MK. KATNB1 is a master regulator of multiple katanin enzymes in male meiosis and haploid germ cell development. Development. 2021;148.

53. Cheung K, Senese S, Kuang J, Bui N, Ongpipattanakul C, Gholkar A, et al. Proteomic analysis of the mammalian Katanin family of microtubule-severing enzymes defines

Katanin p80 subunit B-like 1 (KATNBL1) as a regulator of mammalian Katanin microtubule-severing. Mol Cell Proteomics. 2016;15:1658–1669.
